# Detecting behavioural changes in human movement to inform the spatial scale of interventions against COVID-19

**DOI:** 10.1101/2020.10.26.20219550

**Authors:** Hamish Gibbs, Emily Nightingale, Yang Liu, James Cheshire, Leon Danon, Liam Smeeth, Carl AB Pearson, Chris Grundy, LSHTM CMMID COVID-19 working group, Adam J Kucharski, Rosalind M Eggo

**Affiliations:** Department of Infectious Disease Epidemiology, London School of Hygiene & Tropical Medicine, Keppel Street, London. WC1E 7HT. UK; Department of Global Health and Development, London School of Hygiene & Tropical Medicine, Keppel Street, London. WC1E 7HT. UK; Department of Geography, University College London, Gower St, Bloomsbury, London. WC1E 6BT. UK; Department of Engineering Mathematics, University of Bristol, BS8 1TW. UK; The Alan Turing Institute, British Library, 96 Euston Rd, London NW1 2DB, UK; Population Health Sciences, University of Bristol, BS8, UK; Faculty of Epidemiology and Population Health, London School of Hygiene & Tropical Medicine. UK

## Abstract

**Background:** In 2020, the UK enacted an intensive, nationwide lockdown on March 23 to mitigate transmission of COVID-19. As restrictions began to ease, resurgences in transmission were targeted by geographically-limited interventions of various stringencies. Understanding the spatial scale of networks of human interaction, and how these networks change over time, is critical to inform interventions targeted at the most at-risk areas without unnecessarily restricting areas at low risk of resurgence.

**Methods:** We use detailed human mobility data aggregated from Facebook users to determine how the spatially-explicit network of movements changed before and during the lockdown period, in response to the easing of restrictions, and to the introduction of locally-targeted interventions. We also apply community detection techniques to the weighted, directed network of movements to identify geographically-explicit movement communities and measure the evolution of these community structures through time.

**Findings:** We found that the mobility network became more sparse and the number of mobility communities decreased under the national lockdown, a change that disproportionately affected long distance journeys central to the mobility network. We also found that the community structure of areas in which locally-targeted interventions were implemented following epidemic resurgence did not show reorganization of community structure but did show small decreases in indicators of travel outside of local areas.

**Interpretation:** We propose that communities detected using Facebook or other mobility data be used to assess the impact of spatially-targeted restrictions and may inform policymakers about the spatial extent of human movement patterns in the UK. These data are available in near real-time, allowing quantification of changes in the distribution of the population across the UK, as well as changes in travel patterns to inform our understanding of the impact of geographically-targeted interventions.

**Putting Research Into Context:** *Evidence before this study:* Large-scale intensive interventions in response to the COVID-19 pandemic have been implemented globally, significantly affecting human movement patterns. Mobility data show spatially-explicit network structure, but it is not clear how that structure changed in response to national or locally-targeted interventions.

*Added value of this study:* We used daily mobility data aggregated from Facebook users to quantify changes in the travel network in the UK during the national lockdown, and in response to local interventions. We identified changes in human behaviour in response to interventions and identified the community structure inherent in these networks. This approach to understanding changes in the travel network can help quantify the extent of strongly connected communities of interaction and their relationship to the extent of spatially-explicit interventions.

*Implications of all the available evidence:* We show that spatial mobility data available in near real-time can give information on connectivity that can be used to understand the impact of geographically-targeted interventions and in the future, to inform spatially-targeted intervention strategies.

*Data Sharing:* Data used in this study are available from the Facebook Data for Good Partner Program by application. Code and supplementary information for this paper are available online (https://github.com/hamishgibbs/facebook_mobility_uk), alongside publication.

## Main Text

### Introduction

Fine-scale geographic monitoring of large populations provides a valuable resource for increasing the accuracy and responsiveness of epidemiological modelling, outbreak response, and intervention planning in response to public health emergencies like the COVID-19 pandemic (1–5). Population and mobility datasets collected from the movement of individuals’ mobile phones provide empirical, near-real time metrics of population movement between different geographic regions (6). The COVID-19 pandemic response has benefitted from the availability of new data sources for measuring human movement, aggregated from mobile devices by network providers and popular applications including Google Maps, Apple Maps, Citymapper, and Facebook, and network service providers (7).

Travel and movement behavior during epidemics may change in response to imposed interventions, perceived risk, and due to seasonal activities such as vacations (8,9). During the COVID-19 pandemic, mobility data has been used to assess adherence to movement restrictions (10,11), the impact of movement restrictions on the transmission dynamics of COVID-19 (12–14), and the socioeconomic impacts of large scale movement restrictions (15,16).

In this analysis, we use movement and population data provided by Facebook from March 10 to November 1 2020, which records approximately 15 million daily locations of 4.8 million users (17). We also used population, age, ethnicity, and socioeconomic deprivation data from the UK Office of National Statistics (ONS) to understand the population of users recorded in the movement and population data. We identify changes in travel behavior in response to initially stringent movement restrictions (March to May 2020) and subsequent easing of restrictions, paired with a policy of spatially-targeted interventions in response to local resurgences (May to October, 2020). Using network analytic methods to understand the structure of interconnected communities in the movement network, we trace the evolution of geographic communities through time, comparing them to intervention measures implemented in response to local resurgences.

## Methods

### Facebook Data

Data provided by the Facebook Data for Good partner program (17) uses aggregated and anonymized user data to create a dataset describing user locations in grid cells (Supplemental Figure 1). This data is generated from the population of Facebook users with location services actively enabled.

**Figure 1.**
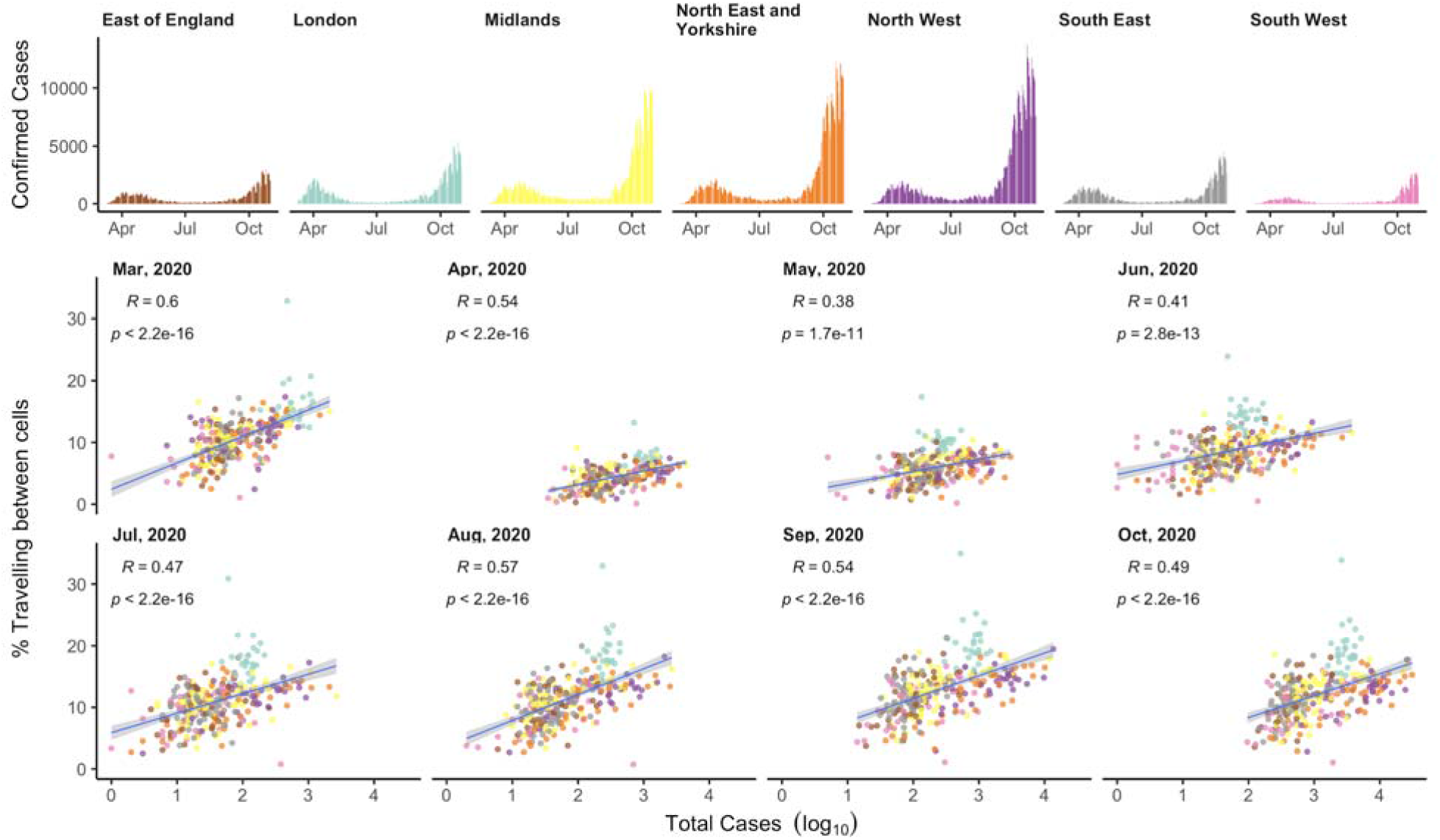
The relationship between movement and COVID-19 cases. a) Daily reported SARS-CoV-2 tests in each NHS region. b) The relationship between the percentage of users travelling outside their cell and the total number of reported SARS-CoV-2 positive tests, by month of the study period. Dots show lower-tier local authorities coloured by their NHS region as in panel a.

We used two datasets; movement and population data, describing users’ modal locations in map cells in sequential 8 hour periods. Population data simply records user locations. In the movement data, sequential user locations define the beginning and end points of journeys between cells. Journeys with fewer than 10 travellers were removed by Facebook prior to data sharing to preserve privacy (Supplemental Figure 2). Any cell that did not record any between-cell journeys with greater than 10 travellers in a given time window was omitted from the dataset, regardless of whether that cell recorded an internal number of users greater than 10.In our network analysis, we constructed a weighted, directed network where nodes were cells, and edge weights were the number of users observed travelling between cells.

**Figure 2.**
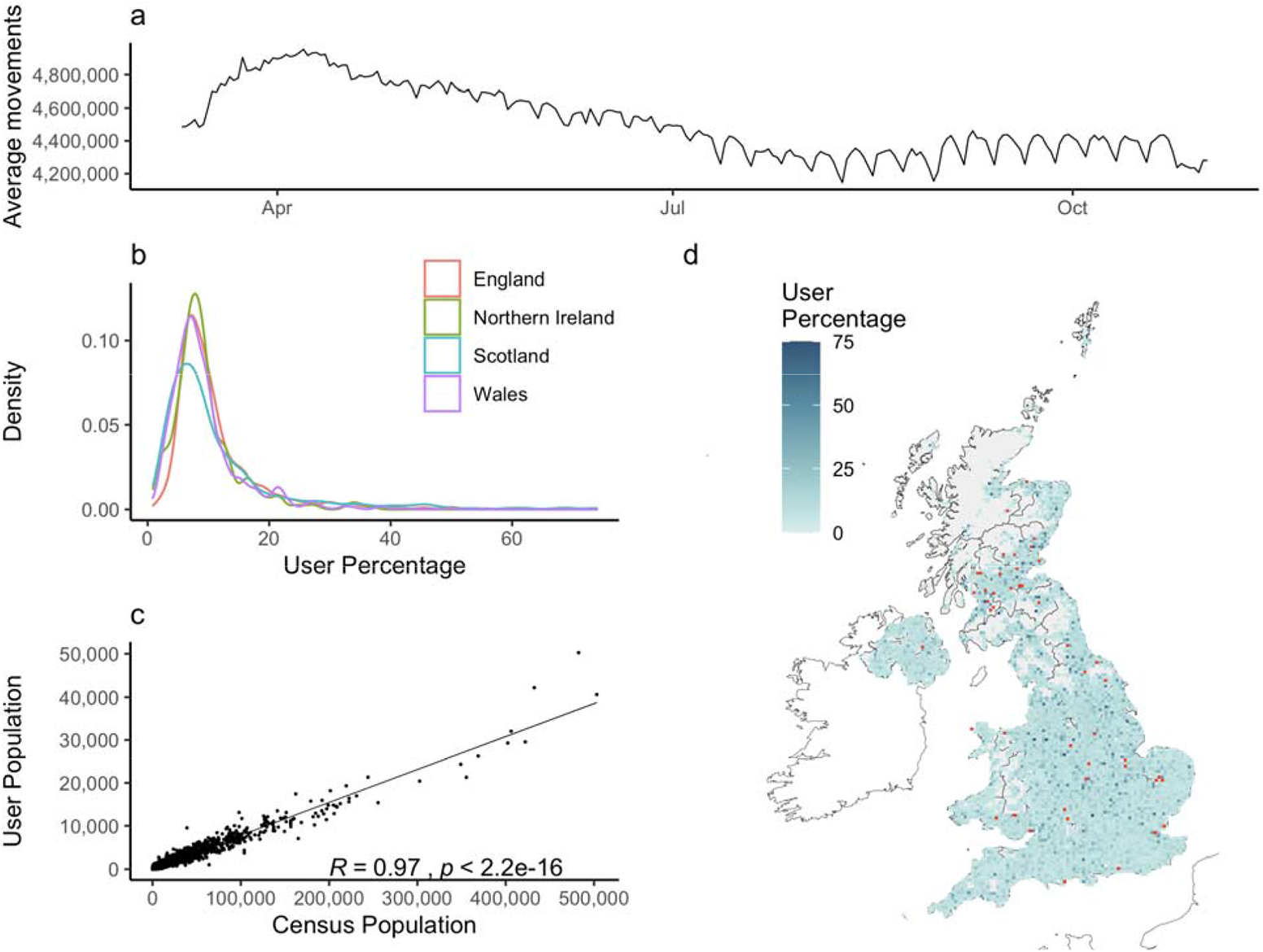
Characteristics of the between-cell Facebook mobility dataset. a) The daily average number of users.b) The probability density functions of the percentage of Facebook users in the census population for cells by country (outliers > 75% removed, see d)). c) The relationship between the number of Facebook users and census population for all cells. d) The spatial distribution of the percentage of Facebook users in the census population for all cells. Grey cells are censored due to low numbers. Red cells are outliers recording >75% of population as users. Note that 12 cells around the town of Swindon are missing due to a data processing error prior to data sharing.

### Bing Maps Tile System

Movement data is referenced to the Bing Maps Tile System, a standard geospatial reference used primarily for serving web maps (18). The system is divided into 23 zoom levels ranging from global level 1, (map scale: 1:295,829,355.45) to detailed level 23, (map scale: 1:70.53). Each Bing Map cell is identified by a “quadkey”, or unique identifier of the zoom level and pixel coordinates of an individual cell. In this analysis, all mobility, population, and other census datasets were referenced to Bing Maps cells. The movement dataset was referenced to cells at zoom level 12 (approximately 4.8 to 6.2 km^2^ in the UK - measured at 60.77° and 50.59° respectively). Facebook population datasets were referenced to cells at zoom level 13 (approximately 2.4 to 3.1 km^2^ in the UK - measured at 60.77° and 50.59° respectively). The ground resolution of cells varies with latitude, with cells at higher latitudes covering a smaller ground area than those at lower latitudes. This distortion results from the distortion inherent in the Web Mercator projection (EPSG:3857) used by the Bing Maps Tile System.

### Demographic information

We compared the age, population, ethnicity, and socioeconomic deprivation of each cell to the population of users recorded in the movement data to identify relationships between the percentage of users and these demographic factors. We extracted these variables from national statistics agencies (Office for National Statistics, Northern Ireland Statistics and Research Agency, Scottish Government, and Welsh Government) and aggregated them to cells. Census variables were referenced to different statistical units by country. In Northern Ireland, census variables were referenced to Super Output areas (SOAs), in England and Wales, Lower Super Output areas (LSOAs), and in Scotland, Data Zones (DZs). Detailed population data was also collected from national statistics agencies, providing a measure of population for Small Areas (Northern Ireland), Output Areas (OA; England and Wales), and Data Zones (Scotland).

Census variables referenced to different national statistical areas were aggregated to align with mobility datasets at zoom level 12. First, we combined the 2011 population weighted centroids of each OA (or equivalent) from the UK Census with 2020 mid-year population estimates in each UK country. We then assigned each OA centroid to the cell it falls within. We then joined 2011-derived census variables (Age, Ethnicity and Socioeconomic Deprivation) to the OA centroids and computed an average of each census variable for each cell, weighted by the OA population estimates. For socioeconomic deprivation data (recorded as ranks) we ranked the weighted average values to create a rank of cells by their population weighted deprivation. Oas are much more granular than cells and therefore nested within them in the majority of cases, minimising the risk of the cells detrimentally intersecting OAs during the demographic assignment.

To assess the correlation between census variables and the proportion of Facebook users in each cell, we computed the Pearson correlation coefficient and two-sided p-values between the proportion of Facebook users in a cell and each census variable.

### Temporal aggregation

Both Facebook movement and population data are recorded in 8 hour intervals. These data display strong and consistent intraday and intraweek patterns. To isolate changes in daily mobility, data collected in 8 hour periods were aggregated to daily periods by taking the sum of the observed number of travellers along a journey for all periods within a day.

### Baseline Population Estimates

To obtain an accurate measurement of the number of users in the Facebook movement and population datasets relative to census population estimates, we used baseline population values computed during the 45 days prior to the creation of the movement and population data collections, from January 29th to March 9th 2020. This baseline population recorded the median number of users in a tile for each daily time window in the reference period. We used the population dataset, rather than movement dataset, because of the higher resolution and reduced impact of censoring on population values. As the population and movement datasets are generated from the same population of users in a given period, we are able to directly compare the user population from the population dataset with that in the movement dataset. To compare user population estimates to movement and census data, we aggregated baseline population estimates from zoom level 13 cells to zoom level 12.

### Community Detection

Community detection methods are algorithms for identifying groups of meaningfully connected vertices in a network. Many methods exist, with various tradeoffs on computational performance, resolution, or other characteristics (19–22). Different community detection methods produce different results because of differences in the network characteristics that they use to define communities. To understand the robustness of the communities detected in this study, we employed two different algorithms, InfoMap and Leiden. InfoMap assesses the movement of a random walker around a network, which identifies communities using the partition of the network that minimizes the description length required to describe the movements (23). The Leiden algorithm maximizes the modularity of different node partitions, identifying the partition for which communities possess stronger connections to community members than to other nodes (24,25).

We compared the effect of the different community detection algorithms, and found that they aligned hierarchically, where the Leiden algorithm identified geographically larger communities. If the communities detected by one method are largely a superset of the communities detected by another, with shared boundaries between the defined communities, this likely represents a differing hierarchical structure, compared to a different interpretation of community structure. We assessed the agreement between community detection methods to understand the stability of detected communities by comparing the proportion of nodes in each community detected using InfoMap with all communities determined using Leiden, and vice versa (Supplemental Figure 9). This comparison allows for the computation of the proportion of shared nodes between both algorithms. The maximum and mean overlap of communities in each algorithm helped to identify the agreement between each method of community detection. In general we found that Leiden detected larger communities, for which the InfoMap communities were (for the most part) sub-communities.

### Community Label Inheritance

The community detection methods used in this study identified communities each day. To track the evolution of communities over the study period, we employed a heuristic approach, assigning the label of a given community identified in a certain time step to that community with the highest number of shared nodes in the following time step (26). When multiple communities in a certain time step “claim” the same community in the following timestep, the community with the closest size to the community in the following timestep “wins” the right to pass its own label to the following timestep.

### COVID-19 Data

We used confirmed COVID-19 cases from the UK Pillar 1 and Pillar 2 testing schemes (27). Pillar 1 is predominantly hospital-based tests including patients and health care workers. Pillar 2 is symptomatic community testing on demand, and represents the bulk of the testing in the UK. Data on the number of confirmed SARS-CoV-2 positive tests by specimen date were available at the Lower Tier Local Authority level (28).

To compare confirmed COVID-19 cases to movement indicators, we measured the total proportion of travellers leaving a grid cell in monthly periods for all cells in England. Cells were then assigned to LTLAs by their maximum areal overlap.

## Results

### Movement patterns observed

Using COVID-19 case data at Lower Tier Local Authority (LTLA) level in England, we identified a consistent association between the proportion of users travelling outside their grid cells and the number of cases in LTLAs per month during the study period (Figure 1, Supplemental Figure 3). While the strength of this association varies at different stages of the pandemic, it provides evidence for the intuitive relationship between increased rates of travel outside of local areas and increased COVID-19 prevalence.

**Figure 3.**
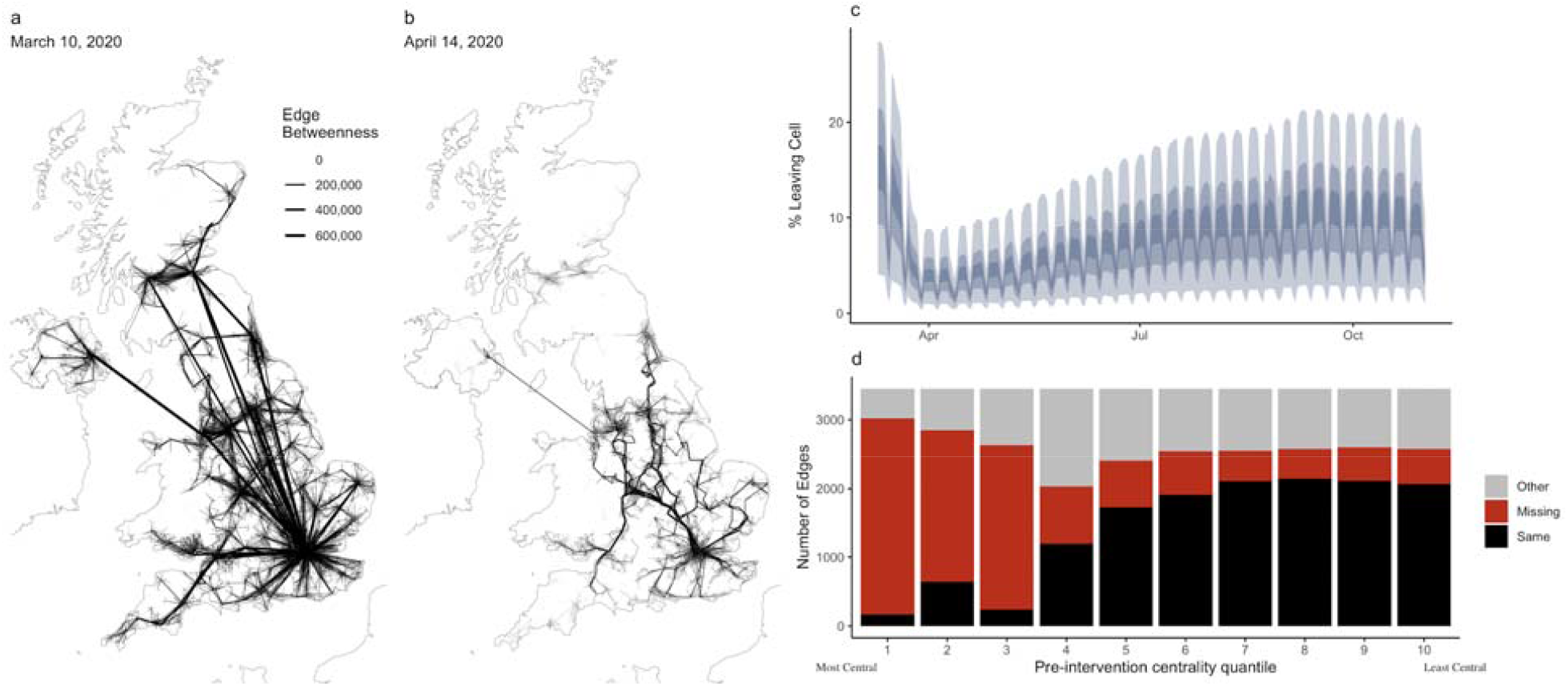
Network structure through time. All network connections on a) Tuesday March 10th 2020, the day of maximum network travel and b) April 14th, 10 days after the announcement of national movement restrictions. The thickness of edges corresponds to their edge betweenness centrality. c) The percentage of users travelling out of their cell through time and d) the likelihood of a journey to remain highly central after the introduction of national interventions. We compare the betweenness quantile for journeys from March 10 to March 23, 2020 (preceding national interventions) and from March 23 to May 10, 2020 (period of national interventions).

To understand the representativeness of Facebook data to the general population, we explored the size of the population of Facebook users included in the movement dataset, and compared this population to 2019 UK census population estimates. The dataset recorded an average of 4.5 million users per 8 hour period, ranging from 5.8 million on March 29th between 4pm and midnight, to 3.7 million on August 9th between midnight and 8am (Figure 2a).

The percentage of Facebook users per cell was comparable in the four nations of the UK (Figure 2b) and was homogeneous across the study area (Figure 2d). The ratio of number of Facebook users and total population was relatively consistent across cells, (Figure 2c). There were no strong associations between the percentage of Facebook users and the average age, percent minority ethnic, population density, or index of multiple deprivation of each cell (Supplemental Figures 4-6).

**Figure 4.**
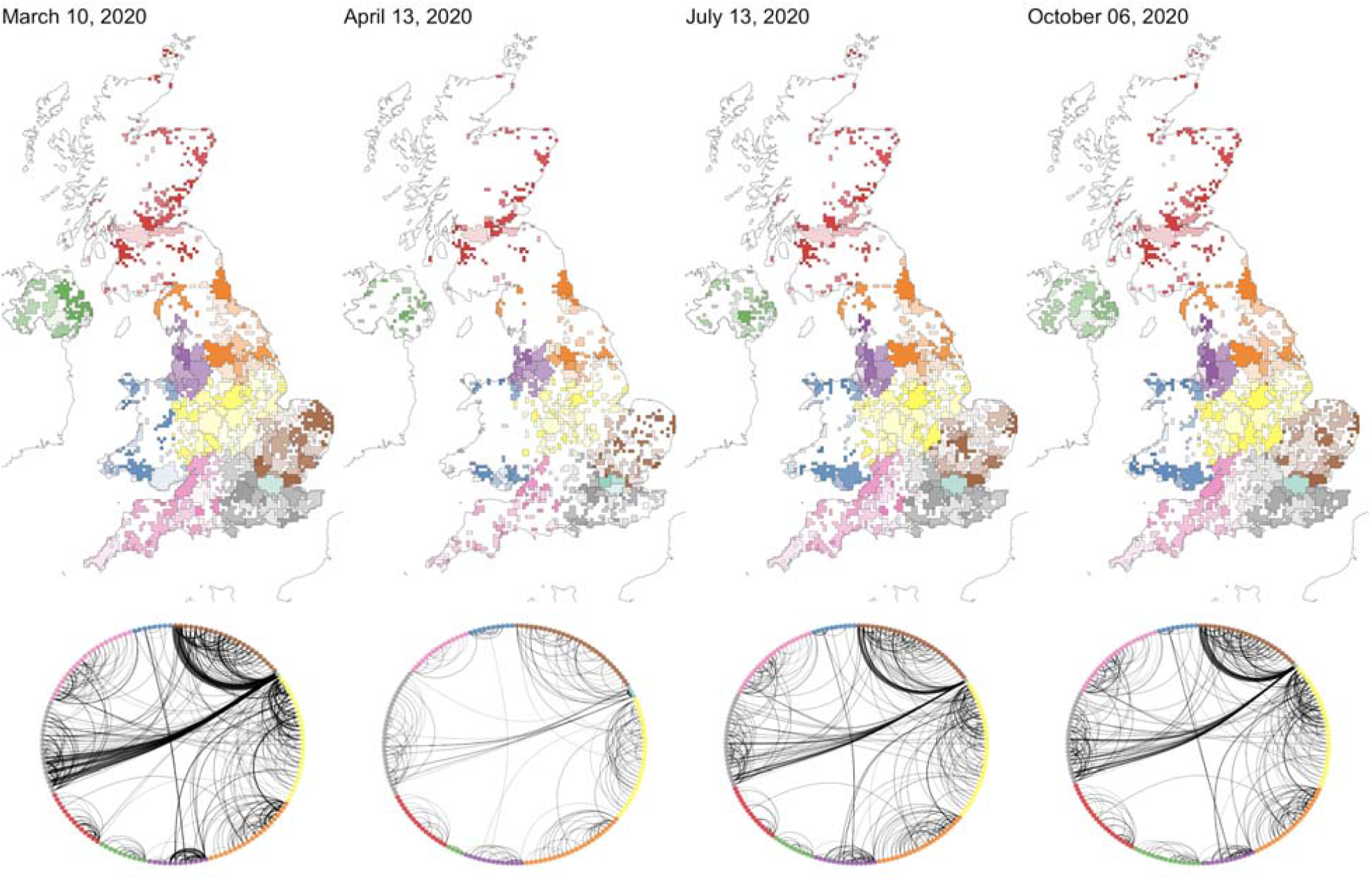
Community detection using the InfoMap algorithm. a) Communities detected on individual days through the time series. Missing tiles which record fewer than 10 users moving have been censored for privacy and appear white. Communities outlined by black lines are coloured by NHS region. Those communities appear in b) in the community network. The number of communities on each day (left to right): 189, 288, 218, 209.

### Network Structure

To quantify how the structure of the overall network changed through time, we computed the edge betweenness centrality of connections between cells, a measure of the relative importance of a given connection in the network (Figure 3a, b). Comparing the period preceding the first national intervention with the period of interventions between March 23, 2020 and May 10, 2020, we compared the likelihood of a journey remaining in the top 10% most central journeys during the intervention period (Figure 3d). During the intervention period we observe a significant decrease in the volume of travel throughout the network, particularly affecting long distance journeys (Supplemental Figure 7). Further, we observe a disproportionate reduction in journeys which were highly central before the introduction of national restrictions, reflecting the reduction of travel along central connections which tend to be highly central in the network (Supplemental Figure 8). Generally, moderately central journeys become highly central after the introduction of nationwide restrictions, while less central journeys remain less central.

Preceding and immediately following the announcement of nationwide travel restrictions on March 23, 2020, we observed a decrease in the volume and distance of between-cell travel, measured by the percentage of users leaving a cell in a given 8-hour period (Figure 3c). While we continued to observe a weekly trend of increased between-tile movements during weekdays, the variance of weekly between-tile journeys decreased during the period of national interventions (Supplemental Figure 9).

We also measured the distance travelled per user in the weighted, directed network, observing a decrease in the overall distance travelled by users during the period of national interventions and a subsequent increase in this distance throughout the summer. As the overall travel in the network decreased, we observed a sharper decrease in the volume of long distance journeys, with most long distance journeys absent from the movement network during national restrictions (Supplemental Figure 7). This decrease reflects both the decreasing volume of long distance travel, and the increased effect of censoring during periods of lower travel volumes.

### Community Detection

We identified geographically-explicit “communities of interaction” in the network of user movements using the InfoMap and Leiden algorithms (Supplemental Figures 10-11). We observed an increase in the number of identified communities and a corresponding decrease in their size following the introduction of nationwide intervention measures on March 23rd, 2020 (Figure 4 and Supplemental Figure 12). The cell-level network also became more sparse as cells were censored from the dataset due to the lower number of journeys between cells. Restrictions were eased incrementally between May and July 2020, during which time we observed an increase in the volume of between-cell movements and an increase in the geographic area and connections between communities.

We found that the most persistent communities existed in some large population centers (Supplemental Figure 14). This reflects both the smaller influence of censoring on higher population cells as well as the continued existence of movement networks, though reduced, around population centres. Persistent communities were identified in Manchester, Newcastle, Glasgow, and Edinburgh, but not in London, which regularly split into more communities on weekends (Supplemental Figure 13-14). We did not find evidence that community stability is associated with population density (Supplemental Figure 15).

By transferring community labels between time windows, we constructed a network of communities in which each community is a node, connected to other communities in a directed network weighted by the number of users travelling between community pairs (Figure 4). In this network, the degree of all nodes decreased after the implementation of nationwide interventions and the overall reduction of between-cell journeys (Supplemental Figure 16). Nonetheless, we did not observe a significant reorganisation in the hierarchy of connections between communities.

### Local Lockdown Extents

After the period of national interventions, the UK introduced local area interventions at differing levels of stringency. The first such intervention was implemented in Leicester on June 30, 2020 in response to a local resurgence (Fig 5b). To understand the impact on the mobility of users, we assessed changes in the volume of travel and network topology before and after introduction.

**Figure 5.**
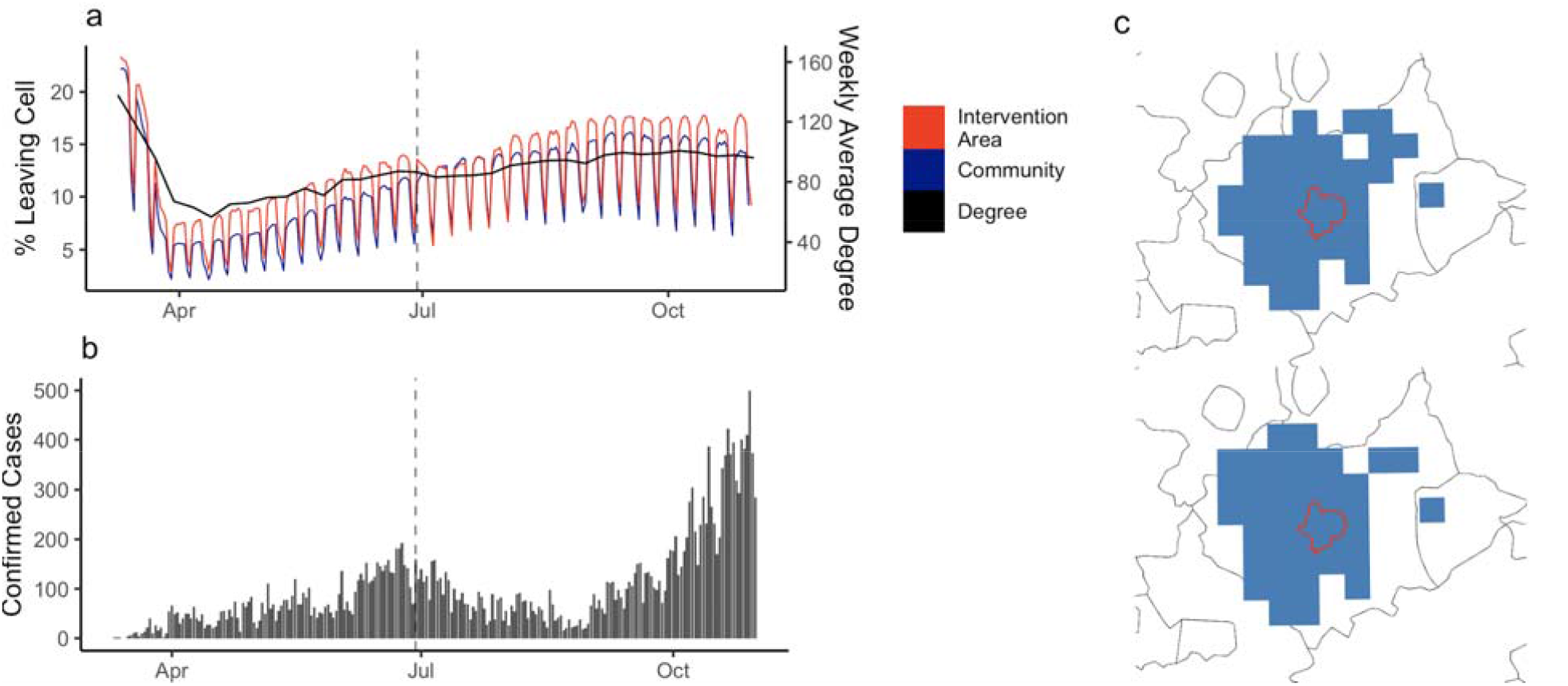
Response to the introduction of local interventions. The daily percentage of users travelling between cells in the intervention area and the connected community, and the weekly average degree of intervention cells (a). Confirmed cases in the intervention area (b). Changes in the community structure before and after the introduction of local intervention (c).

We measured the connection between cells overlapping areas of local interventions in four areas: Leicester (Fig 5), Manchester, the North West, and the North East (Supplementary Figures 17-19) to assess the impact on volume of travel and the isolation of intervention areas from the broader UK movement network. We found that, while movement indicators did decrease, particularly in Leicester, the response was smaller than during the first national lockdown (Figure 5a).

Motivated by the need to identify communities associated with epidemic resurgences and responses to reactive interventions, we compared the extent and date of local interventions with the spatial extent and temporal persistence of network communities (Figure 5c). We found that network communities remained relatively stable after the introduction of intervention measures in all areas, with some peripheral changes to movement communities in Manchester, the North West, and North East (Supplementary Figures 17-19).

From July 2020 onwards, the geographic extent of local area interventions was in closer agreement with movement communities, particularly in Manchester (Supplemental Figure 17). Some interventions also spanned multiple movement communities, as in the North West and

North East of England (Supplemental Figures 18-19). Early local intervention measures at limited spatial extents may not have fully encompassed the area of transmission resurgence, however UK policy changed over time, beginning to enforce collaborative local area interventions comprising multiple Local Authorities. Additionally, movement communities evolve over time, and have the potential to shift following local area interventions, requiring an understanding of real-time patterns of movement to monitor the appropriateness of a given measure.

## Discussion

This study used a large, anonymized movement dataset to quantify changes in the UK movement network and assessed how geographic communities were affected by interventions. Using movement communities, we can identify strongly connected areas that can be used to inform the spatial scale of geographically-targeted interventions to respond to resurgences of COVID-19. We also explored the structure of the UK travel network through the pandemic, identifying variations in the central connections between population centres and changing travel patterns in response to the introduction of public health measures.

Gridded mobility datasets such as those used in this study provide granular, near real-time information about the movement patterns of a large sample of the UK population. While these datasets could usefully inform epidemic responses (15,29–31), there remain questions about the generalizability of the movement recorded in these datasets to the movements of the overall population (32,33). The privacy preserving structure of the Facebook movement dataset means that low frequency journeys are not recorded, precise locations are replaced by grid cell references, and data is provided in a uniform-area grid cells which vary in population size. Ideally, multiple mobility datasets should be analysed to improve the interpretation of changes in mobility indicators.

In response to the nationwide lockdown introduced March 23rd, the UK mobility network changed drastically. The movement network became more sparse, with reductions in travel volume and distance. These changes disproportionately affected long distance journeys and highly central journeys, important connections which integrated geographically distant areas in the broader UK travel network.

Using a network analysis of mobility data, we identified geographically distinct communities with strong interconnections that are relevant to policy responses focused on limiting transmission in response to geographically-limited disease resurgences. These communities provide empirical boundaries that can inform policy responses. We found that these communities were stable around some population centres, and that the spatial extent and number of communities changed during the implementation of public health interventions, particularly during the national intervention between March and May 2020. As restrictions evolve in the UK, the boundaries and size of communities may change, a response to new patterns of travel, home-working, and commuting.

In response to disease resurgences in a particular area, determining the geographic extent of reactive interventions should be driven by areas at risk of increased transmission, which may not intersect with administrative boundaries. We found that while communities tended to stabilise around settlements, there was disagreement between the extent of these communities and the boundaries at which local area interventions have been introduced in the UK. While we do not advocate the sole use of movement communities to delineate the extent of intervention measures, these communities provide valuable information in near-real time about the extent of typical patterns of travel, their temporal variations, and “catchment areas” of movement around a given area.

There are several caveats to the methods of community detection used in this study, as the extent of communities could be influenced by the level of aggregation of the Facebook mobility data, and cells were assigned to a single community each day. While we conducted a sensitivity analysis using two methods for identifying communities, there are a wide variety of community detection algorithms which emphasize different aspects of network structure. Questions also remain about the general reliability of community detection methods, which have been developed on well understood network structures, when applied to real-world networks (20). The effect of local area interventions on travel depends on the specifics of each intervention and their stringency. Additionally, interventions occur at multiple spatial scales, and across overlapping time periods. For example, in the UK, national interventions coincide with local interventions, and each may contribute differently to changes in movement behaviour.

## Conclusion

Data-driven approaches using mobility data can help to quantify patterns of travel and inform geographically-targeted public health interventions.

## Data Availability

Available on GitHub.

## Ethics Approval

This research was approved by the LSHTM Observational Research Ethics Committee (ref 16834-1).

## Author Contributions

RME conceived the project, and developed the methods and analytical plan with HG, CABP, EN, and YL. JC and LS contributed to analysis of census variables. CABP and LD contributed to community detection methods and analysis of the methods. JC and CG contributed to analysis of spatial data. YL and EN contributed to statistical analysis of the mobility, census, and case data. AJK acquired the data and contributed to the analysis and understanding of Facebook data. HG wrote and implemented the majority of the code, and made the figures, with input from all authors. HG and RME wrote the first draft, to which all authors contributed. All authors interpreted the findings and were involved in the analysis and decision to publish.

Facebook had no role in study design, the analysis, or decision to publish.

## Acknowledgements

The following funding sources are acknowledged as providing funding for the named authors. This research was partly funded by the Bill & Melinda Gates Foundation (INV-003174: YL; NTD Modelling Consortium OPP1184344: CABP; OPP1183986: ESN). DFID/Wellcome Trust (Epidemic Preparedness Coronavirus research programme 221303/Z/20/Z: CABP). This project has received funding from the European Union’s Horizon 2020 research and innovation programme - project EpiPose (101003688: YL). HDR UK (MR/S003975/1: RME). UK DHSC/UK Aid/NIHR (PR-OD-1017-20001: HPG). UK MRC (MC_PC_19065: RME, YL). Wellcome Trust (206250/Z/17/Z: AJK), UK MRC (MC_PC_19067, MR/V038613/1 LD), UK EPSRC (EP/V051555/1: LD), The Alan Turing Institute under the EPSRC (grant EP/N510129/1: LD). This research was partly funded by the National Institute for Health Research (NIHR) using UK Aid from the UK Government to support global health research. The views expressed in this publication are those of the author(s) and not necessarily those of the NIHR or the UK Department of Health and Social Care (16/137/109: YL; NIHR200908: RME).

The following authors were part of the Centre for Mathematical Modelling of Infectious Disease COVID-19 working group. Each contributed in processing, cleaning and interpretation of data, interpreted findings, contributed to the manuscript, and approved the work for publication: Sam Abbott, Kaja Abbas, Kiesha Prem, Sebastian Funk, Jon C Emery, Georgia R Gore-Langton, Fiona Yueqian Sun, Arminder K Deol, Alicia Showering, Nicholas G. Davies, Nikos I Bosse, Samuel Clifford, Anna M Foss, Graham Medley, C Julian Villabona-Arenas, Timothy W Russell, Amy Gimma, W John Edmunds, Gwenan M Knight, Yung-Wai Desmond Chan, Yalda Jafari, Quentin J Leclerc, Rein M G J Houben, Akira Endo, Sophie R Meakin, Petra Klepac, Joel Hellewell, Naomi R Waterlow, Kevin van Zandvoort, Christopher I Jarvis, Rachel Lowe, Matthew Quaife, Charlie Diamond, Megan Auzenbergs, Simon R Procter, Rosanna C Barnard, Oliver Brady, Katherine E. Atkins, Katharine Sherratt, Thibaut Jombart, Stéphane Hué, Kathleen O’Reilly, Jack Williams, David Simons, Stefan Flasche, Mark Jit, James D Munday, Billy J Quilty, Frank G Sandmann, Damien C Tully, James W Rudge, Alicia Rosello.

The following funding sources are acknowledged as providing funding for the working group authors. Alan Turing Institute (AE). BBSRC LIDO (BB/M009513/1: DS). This research was partly funded by the Bill & Melinda Gates Foundation (INV-001754: MQ; INV-003174: KP, MJ; NTD Modelling Consortium OPP1184344: GFM; OPP1180644: SRP; OPP1191821: KO’R, MA). BMGF (OPP1157270: KA). DFID/Wellcome Trust (Epidemic Preparedness Coronavirus research programme 221303/Z/20/Z: KvZ). DTRA (HDTRA1-18-1-0051: JWR). Elrha R2HC/UK DFID/Wellcome Trust/This research was partly funded by the National Institute for Health Research (NIHR) using UK aid from the UK Government to support global health research. The views expressed in this publication are those of the author(s) and not necessarily those of the NIHR or the UK Department of Health and Social Care (KvZ). ERC Starting Grant (#757699: JCE, MQ, RMGJH). This project has received funding from the European Union’s Horizon 2020 research and innovation programme - project EpiPose (101003688: KP, MJ, PK, RCB, WJE). This research was partly funded by the Global Challenges Research Fund (GCRF) project ‘RECAP’ managed through RCUK and ESRC (ES/P010873/1: AG, CIJ, TJ). MRC (MR/N013638/1: NRW). Nakajima Foundation (AE). NIHR (16/136/46: BJQ; 16/137/109: BJQ, CD, FYS, MJ; Health Protection Research Unit for Immunisation NIHR200929: NGD; Health Protection Research Unit for Modelling Methodology HPRU-2012-10096: TJ; NIHR200929: FGS, MJ; PR-OD-1017-20002: AR, WJE). Royal Society (Dorothy Hodgkin Fellowship: RL; RP\EA\180004: PK). UK MRC (LID DTP MR/N013638/1: GRGL, QJL; MC_PC_19065: AG, NGD, SC, TJ, WJE; MR/P014658/1: GMK). Authors of this research receive funding from UK Public Health Rapid Support Team funded by the United Kingdom Department of Health and Social Care (TJ). Wellcome Trust (206250/Z/17/Z: TWR; 206471/Z/17/Z: OJB; 208812/Z/17/Z: SC, S Flasche; 210758/Z/18/Z: JDM, JH, KS, NIB, SA, SFunk, SRM). No funding (AKD, AMF, AS, CJVA, DCT, JW, KEA, SH, YJ, YWDC).

