## Supplementary Information for "Detecting behavioural changes in human movement to inform the spatial scale of interventions against COVID-19"

**Affiliations:**

^4^Department of Computer Science, University of Exeter, Stocker Rd, Exeter. EX4 4PY. UK

### Data Representativeness

To understand the representativeness of the Facebook Movement and Population datasets, we compared the percentage of the population recorded by the Facebook movement dataset with UK census variables aggregated to the same spatial unit as the Facebook cell movement dataset. Each country of the UK (England, Scotland, Wales, Northern Ireland) was compared independently because of differences in the collection of the census variables.

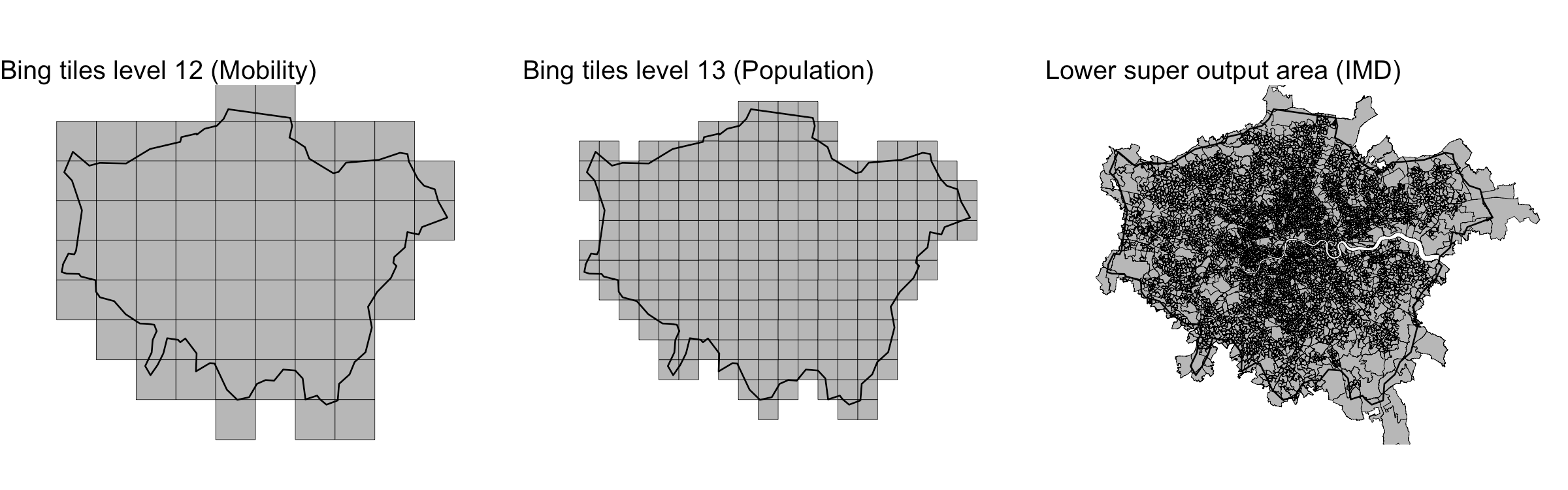

**Supplemental Figure 1. An overview of different geometries used in this study, intersecting London.** a) Zoom level 12 tiles, b) Zoom level 13 tiles, c) Lower Super Output Areas.

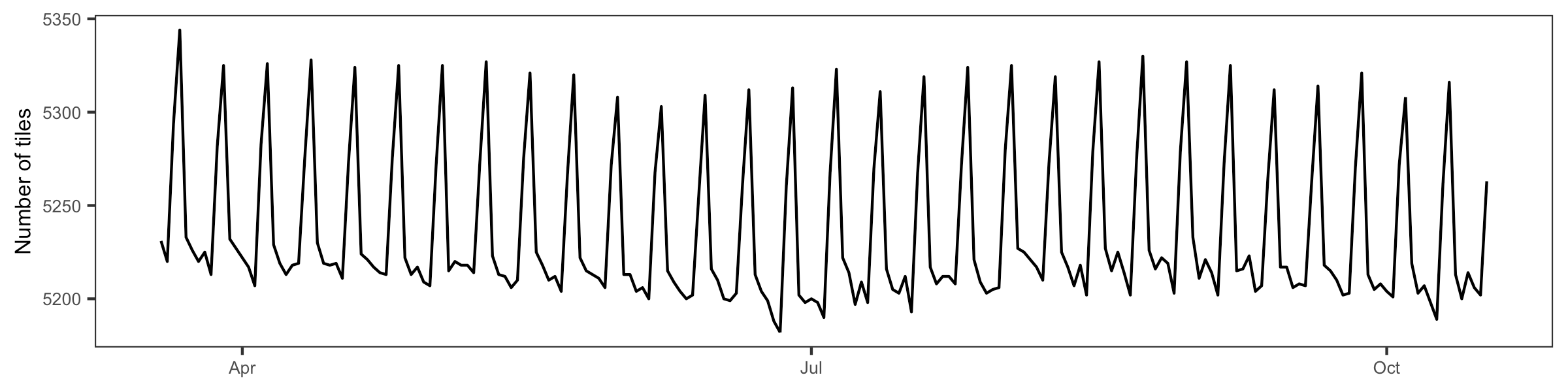

**Supplemental Figure 2. The number of cells included in the dataset.** Cells recording fewer than 10 persons moving between cells along any connection are censored from the dataset to preserve user privacy.

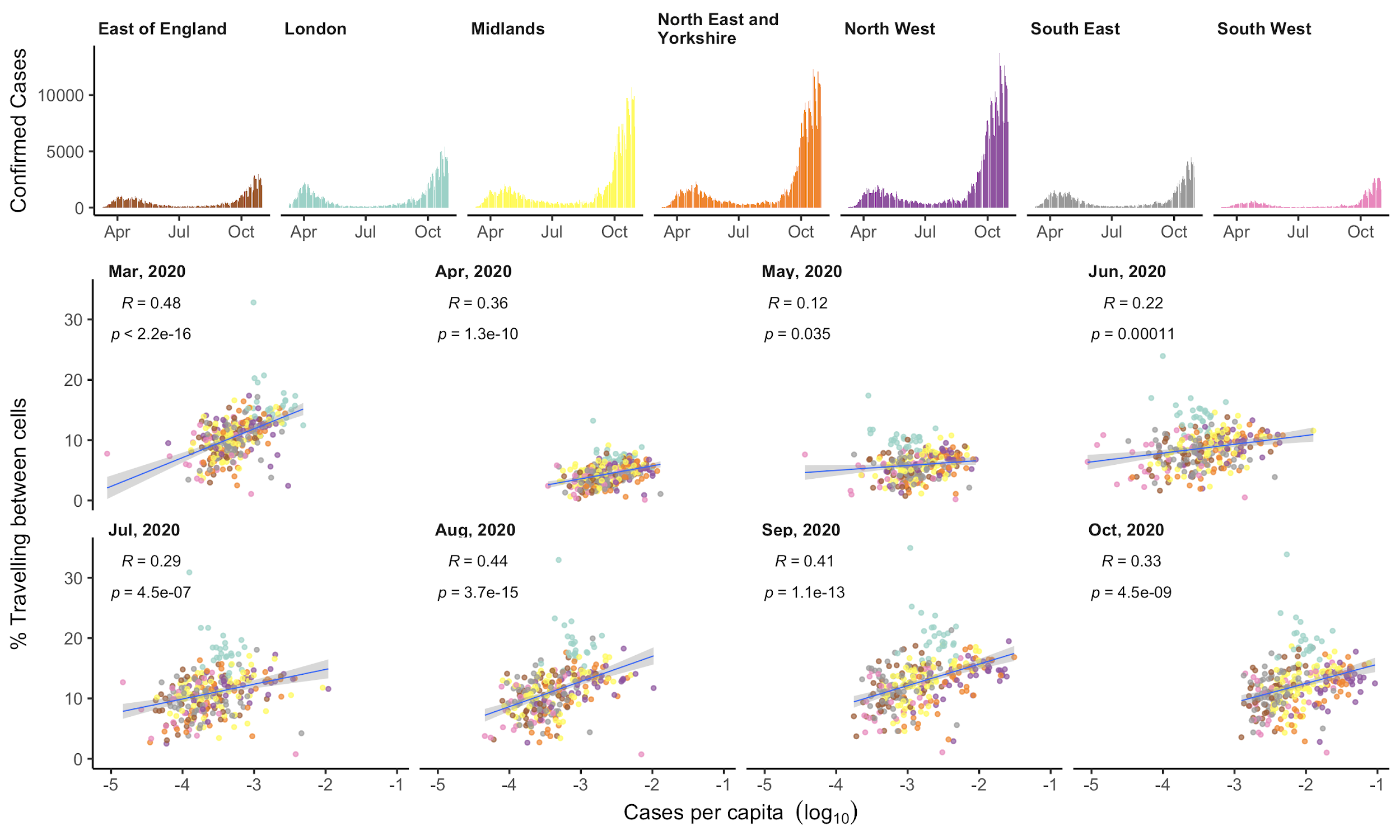

**Supplemental Figure 3. The relationship between movement and COVID-19 cases per capita.** a) Daily reported SARS-CoV-2 tests in each NHS region. b) The relationship between the percentage of users travelling outside their cell and the total number of reported SARS-CoV-2 positive tests per capita, by month of the study period. Dots show lower-tier local-authorities coloured by their NHS region as in panel a.

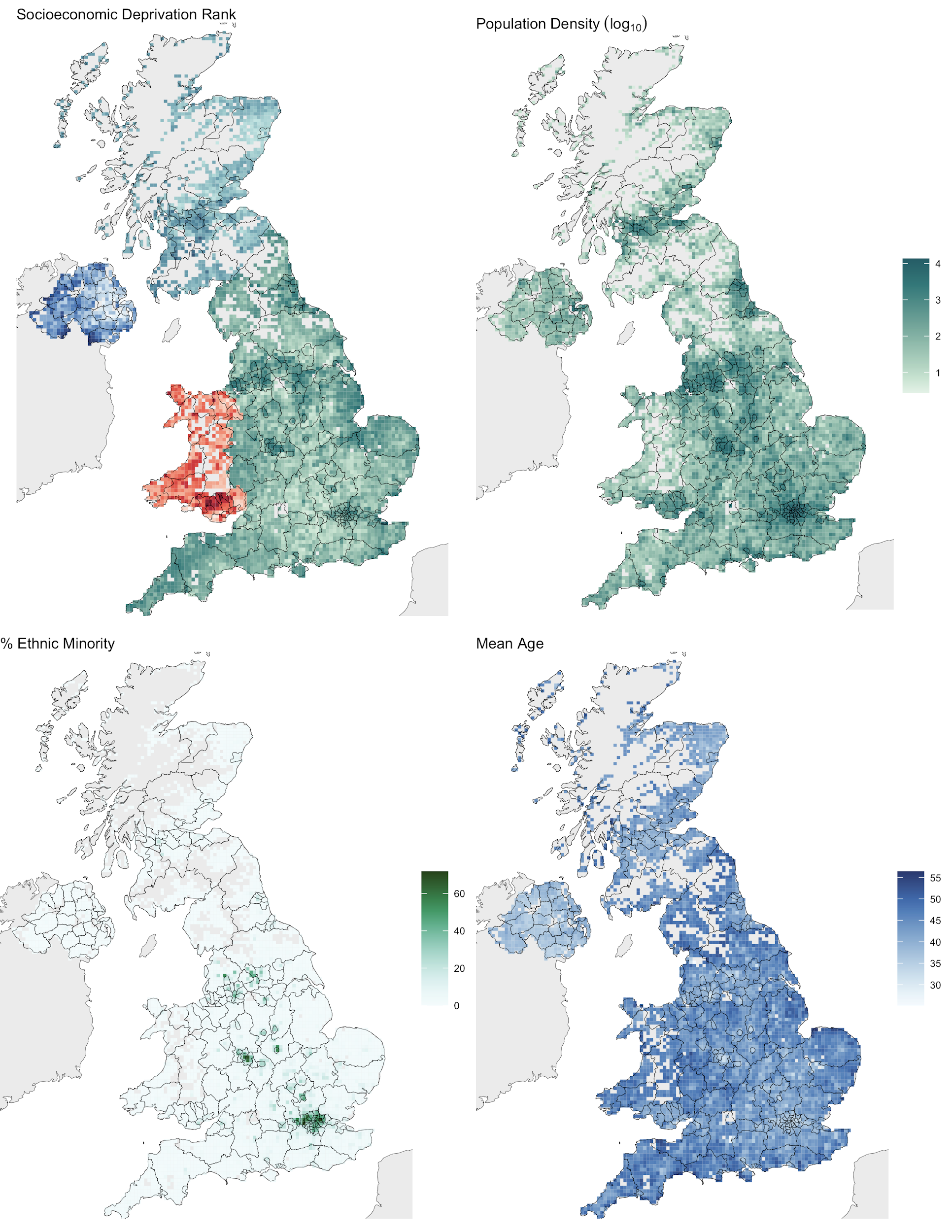

**Supplemental Figure 4. The cell-level geographic distribution of the four census variables used in the analysis.** In each case, a white cell means the data were missing from the Facebook mobility data and so are not displayed here. In most cases this is due to censoring of low numbers, except for the small discontinuity around Swindon, mentioned in the Main Text. a) Socioeconomic deprivation rank. Each country has a different colour because the measure of socioeconomic deprivation is different in each country. In each case, the darker shade is higher deprivation. b) Population density per cell (log scale). c) Percentage of the population self-identifying as any other ethnicity than “Any white background”. d) Mean age of the population resident in each cell.

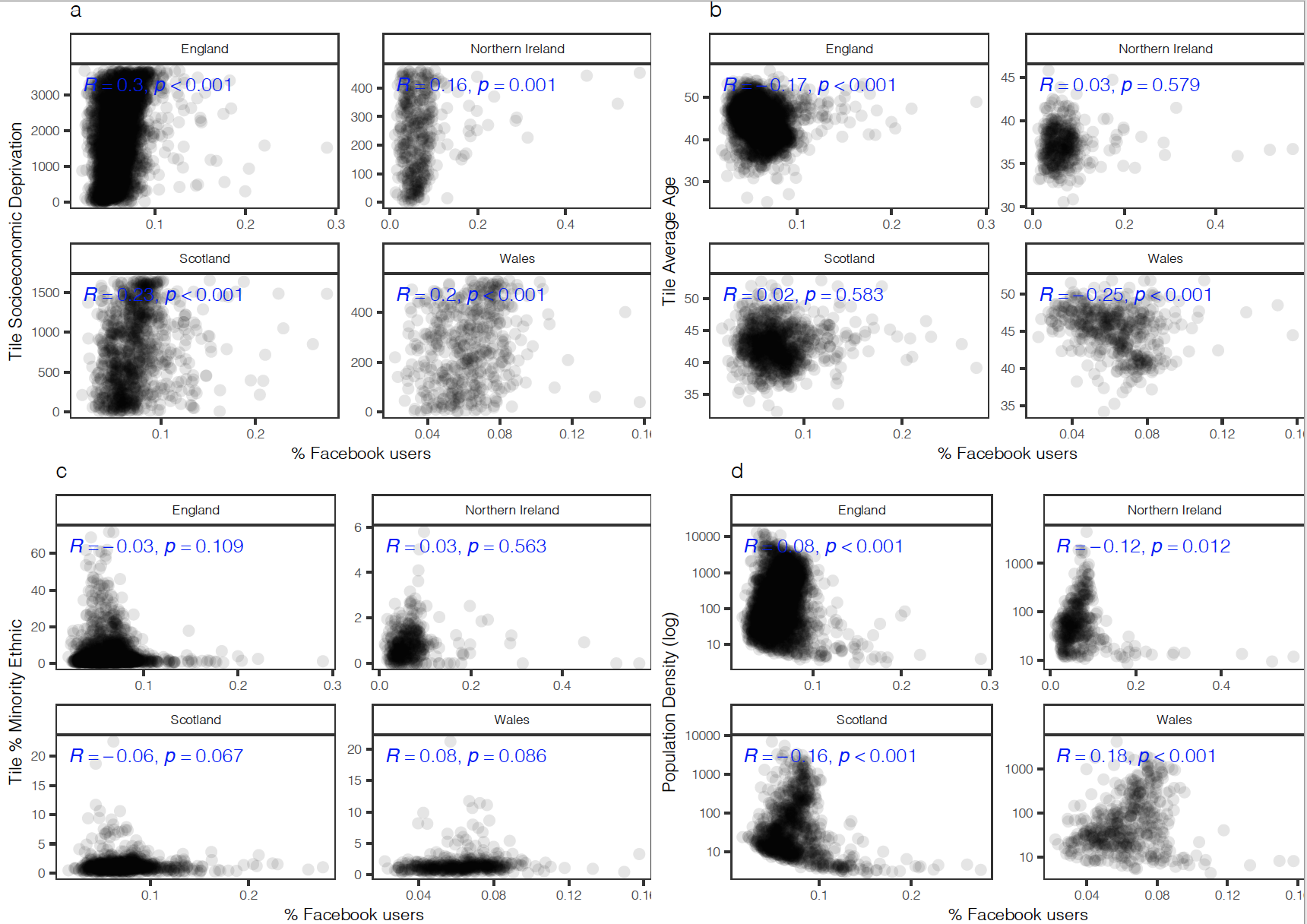

**Supplemental Figure 5. A comparison of the percentage of Facebook users and census variables at the cell level.** a) socioeconomic deprivation rank, b) mean age, c) percent minority ethnic, and d) population. Variables were aggregated from mid-level census geographies for each country. The mean value of each variable was assigned to intersecting tiles, weighted by small area population estimates. Correlation is shown on the panel as the Pearson correlation coefficient (*R*) and two-sided p values.

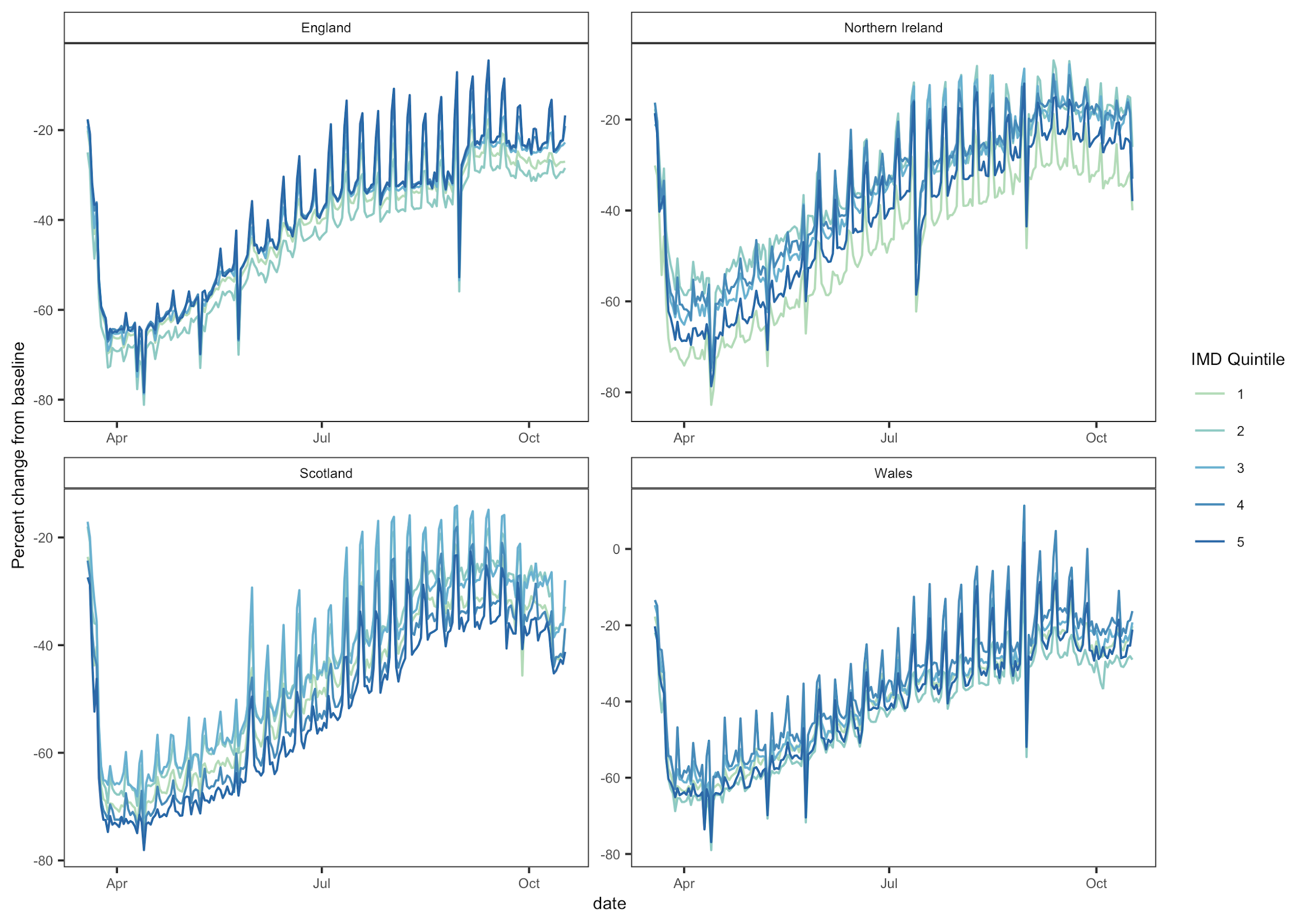

**Supplemental Figure 6. A comparison of movement by IMD quintile.** Percent change from baseline for movement between cells by IMD quintiles in each country. IMD data was aggregated to cell level and weighted by small area population estimates. IMD quintiles range from 1 (most deprived) to 5 (least deprived).

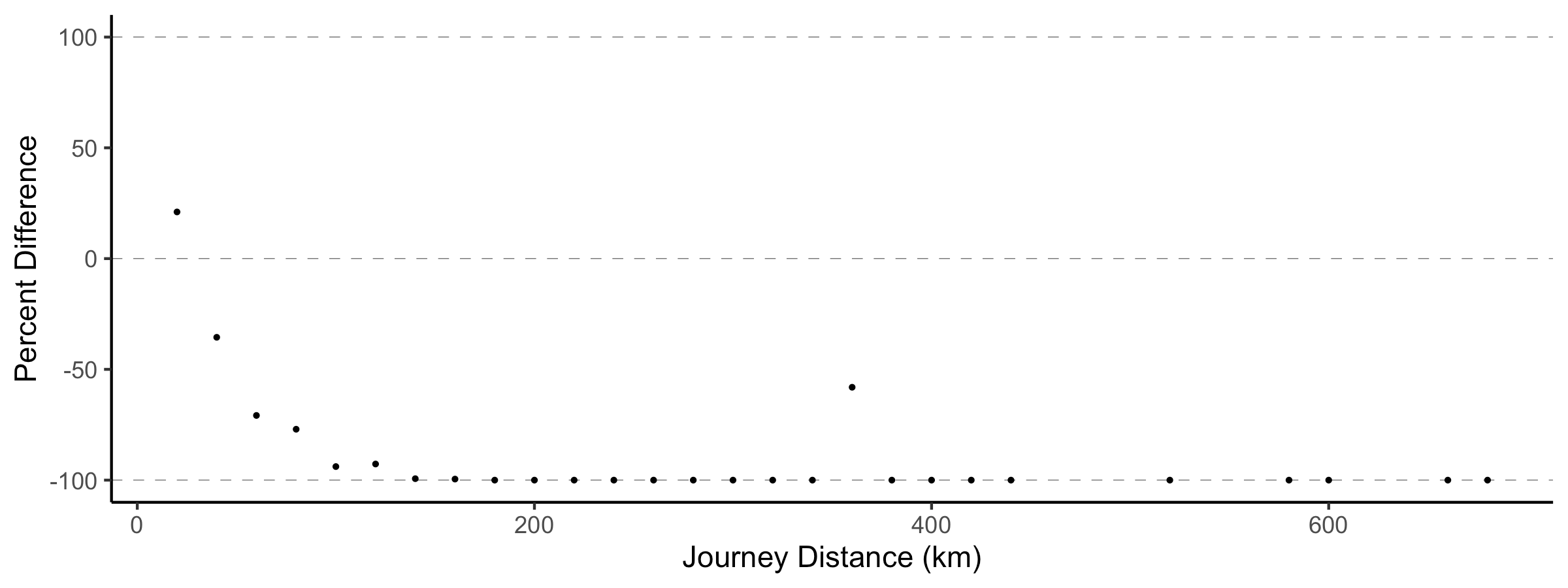

**Supplemental Figure 7. Reduction of travel by distance during national interventions.** We observe a sharp decrease in the volume of travel along long-distance journeys during the period of national interventions. Most long-distance journeys are removed from the dataset entirely as a result of decreased travel and the impact of censoring to preserve privacy. Note that the longest remaining connection is between Liverpool and Belfast.

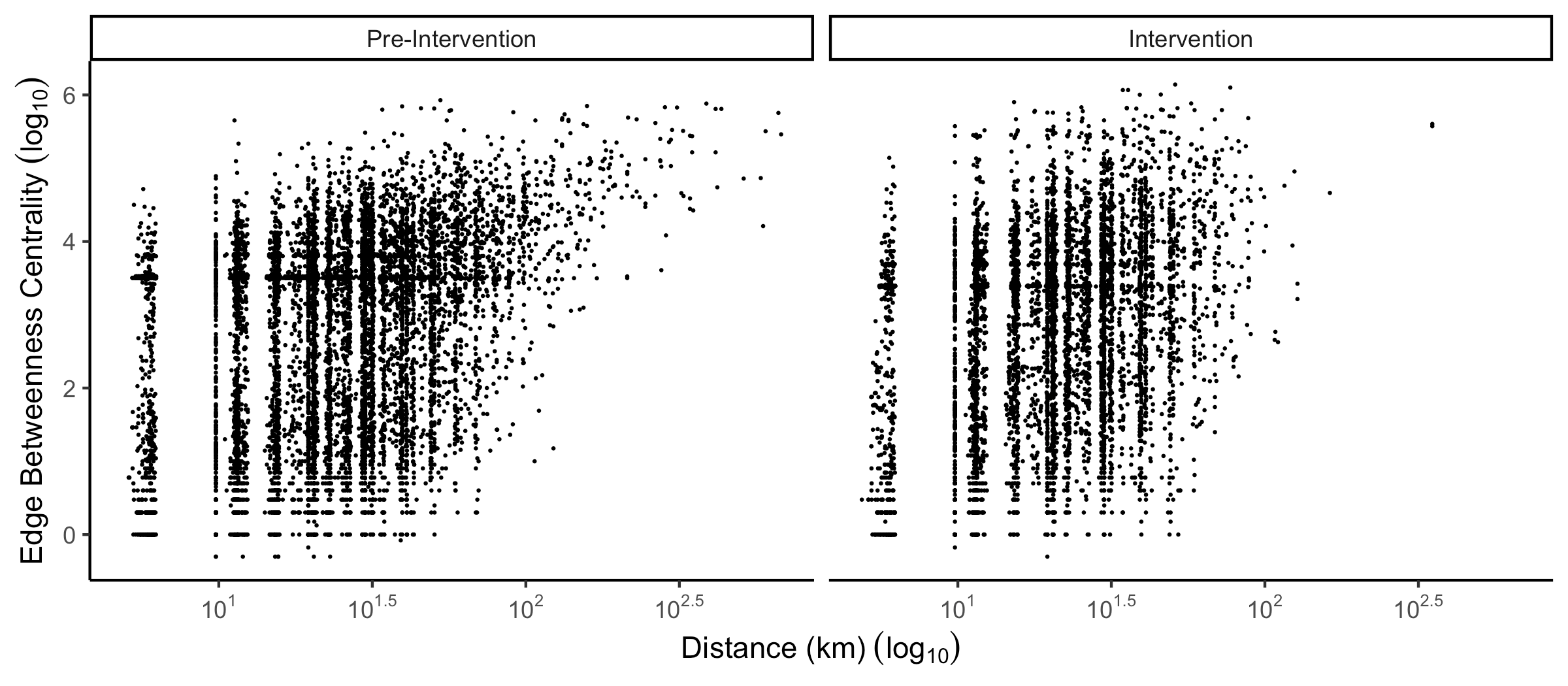

**Supplemental Figure 8. The relationship between edge betweenness centrality and distance.** The relationship between edge betweenness centrality and journey distance before and after the introduction of national interventions. Long, central journeys are significantly reduced in the intervention travel network.

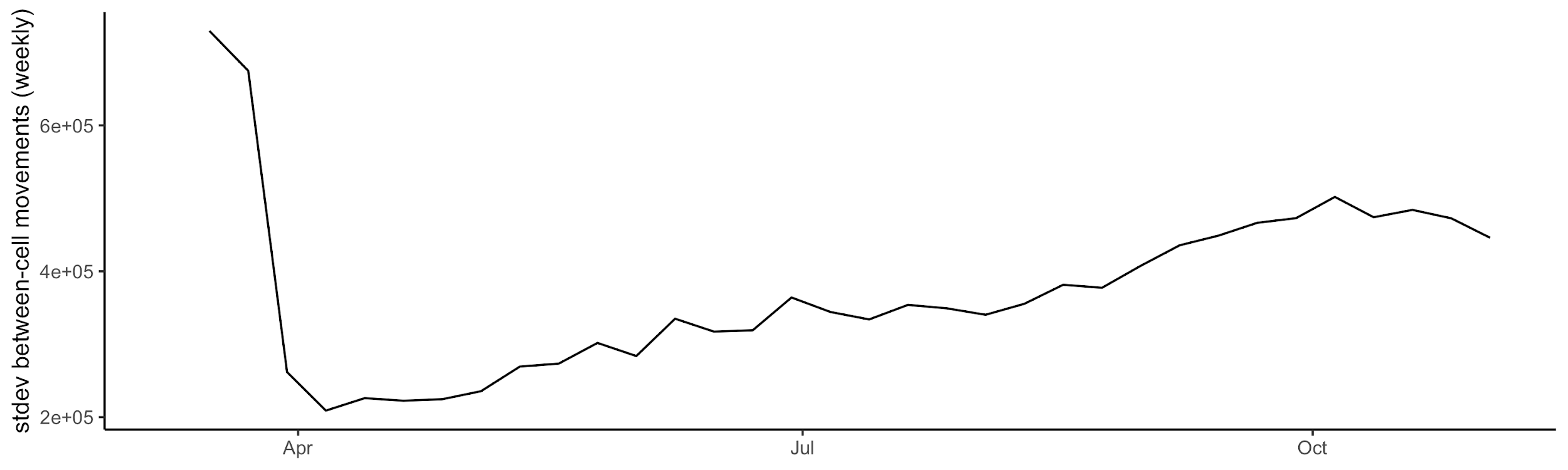

**Supplemental Figure 9. Weekly variance of between-cell movement.** The standard deviation of between cell movements through time. Decreased variance indicated smaller differences in daily between-cell travel measurements per week. This reflects a reduction in the weekly pattern of between-cell movements.

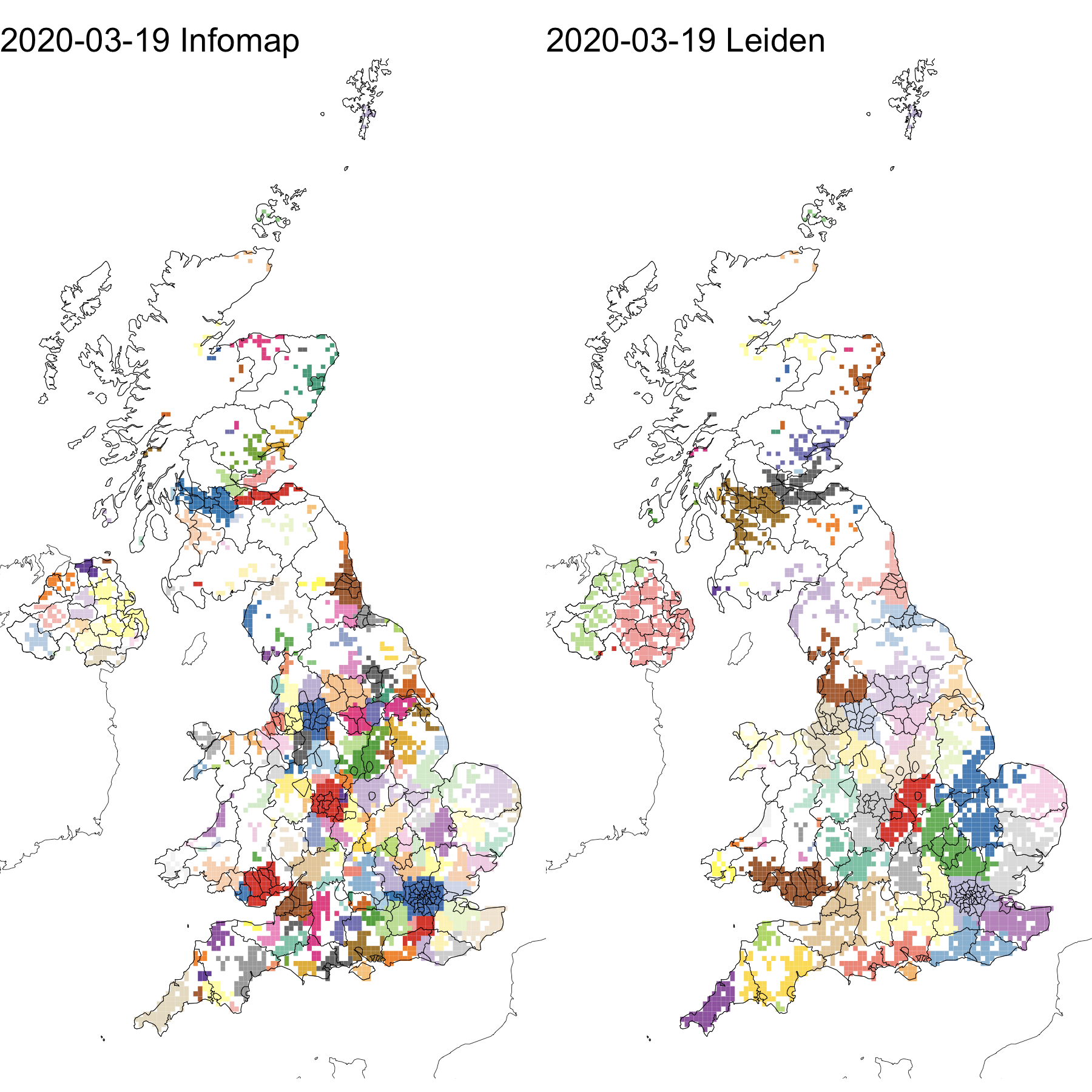

**Supplemental Figure 10. Spatial comparison of community detection algorithms.** The extent of communities detected by InfoMap (a) and Leiden (b) on March 19th. Leiden communities are largely a superset of communities detected by Infomap, indicating the detection of a different hierarchical structure, but an agreement of community boundaries between the two algorithms.

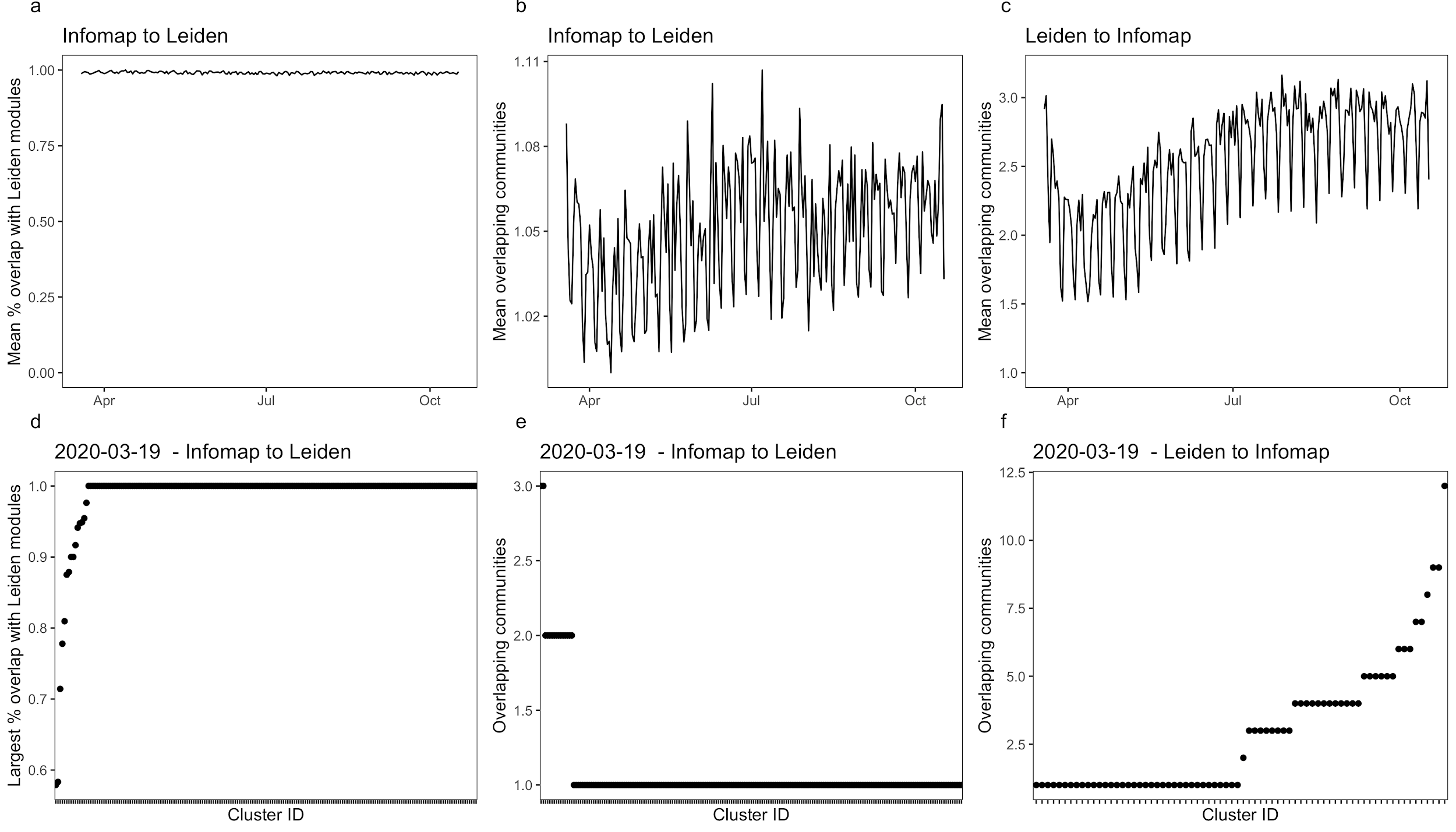

**Supplemental Figure 11. Comparing community detection algorithms.** A comparison of the spatial intersection between communities detected by the Infomap and Leiden algorithms over time (a) and on March 19th (b), the first day of the time series. Each Infomap community was compared to all Leiden communities and each Leiden community was compared to all InfoMap communities. The percent overlap between communities and the number of intersecting communities were compared. The mean values of each metric were taken to compare changes over time.

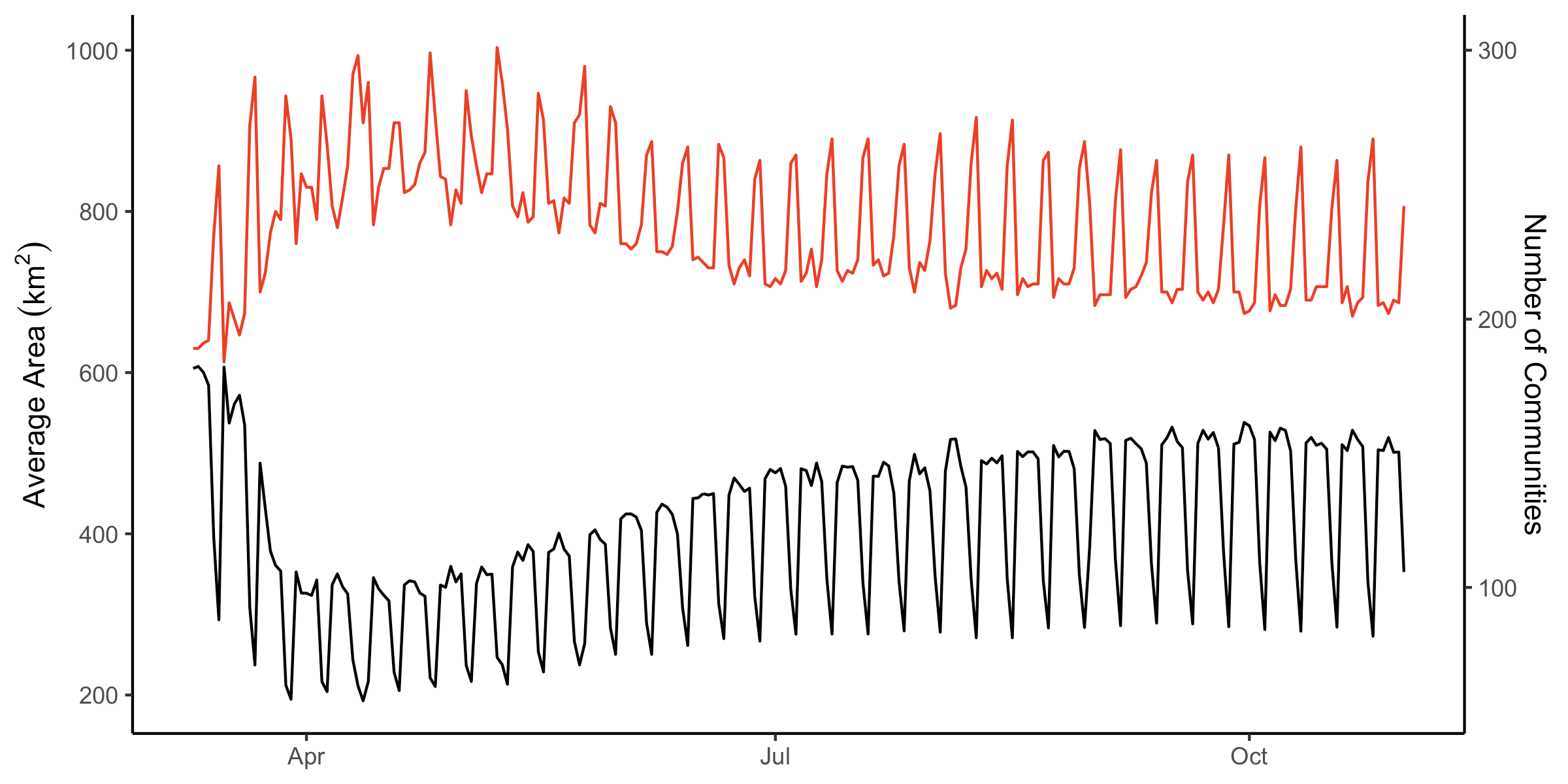

**Supplemental Figure 12. Characteristics of InfoMap communities.** The number (red) and area(black) of InfoMap communities through time. An increase in the number and corresponding decrease in the area of communities reflects the more local patterns of travel during national interventions.

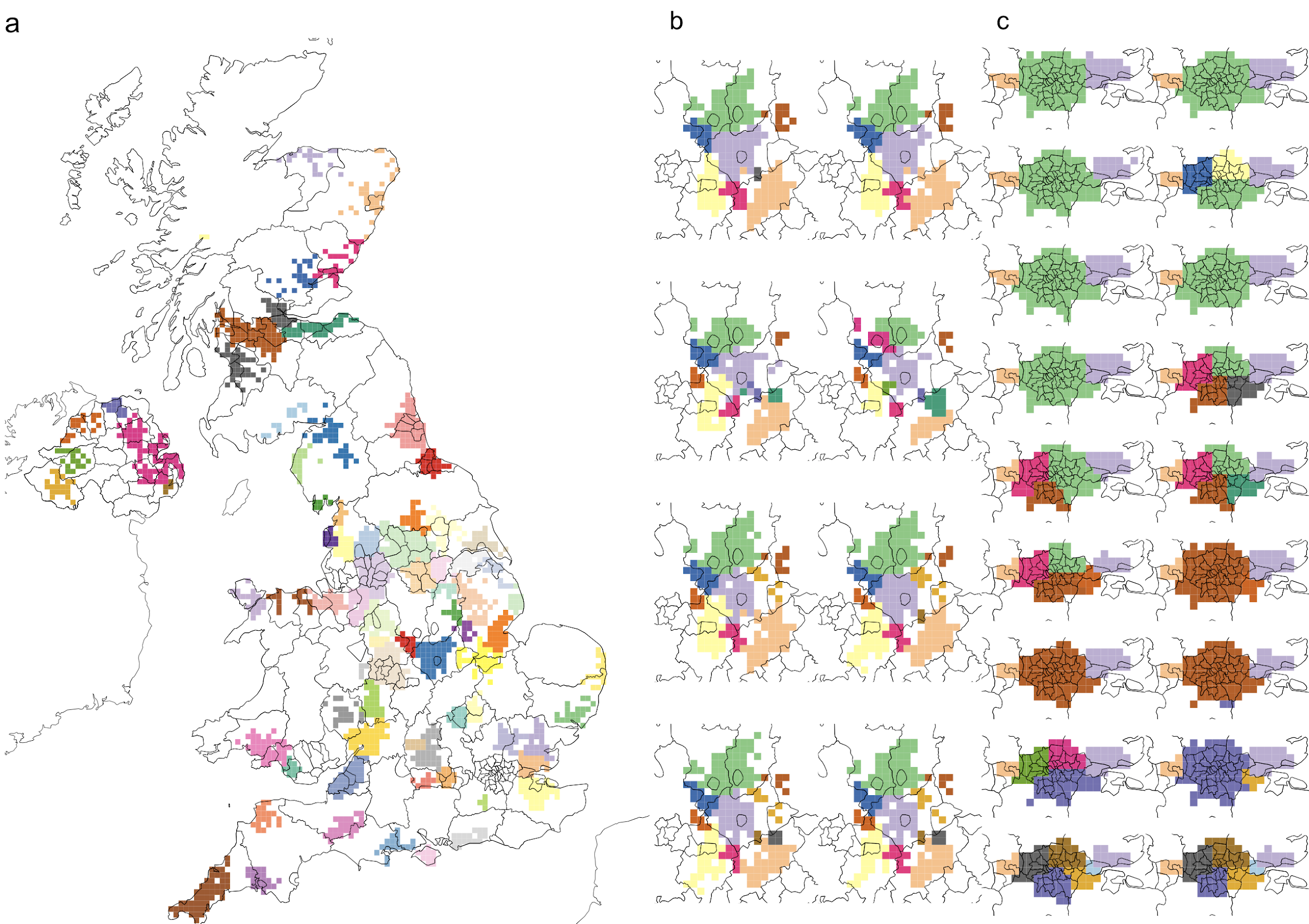

**Supplemental Figure 13. Community persistence in the dataset.** a) The most persistent communities, those that existed throughout the timeseries, as on March 19th, 2020. b) Community membership from March 19th to March 26th, 2020 in Leicestershire, and c) community membership from March 19th to April 5th, 2020 in London.

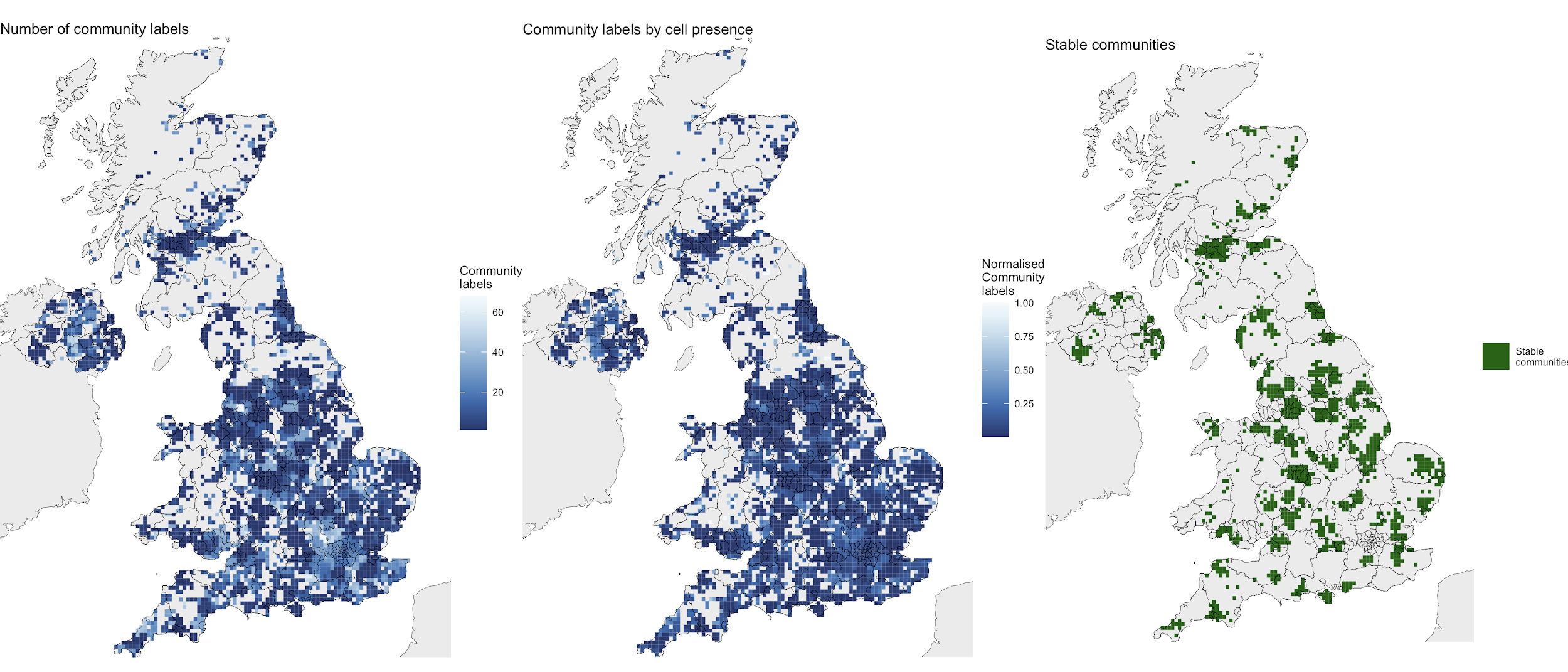

**Supplemental Figure 14. Persistence of communities.** a) the total number of community labels that each cell has had (i.e. number of communities that the cell has ever been in) during the study period. The darkest shade indicates that a cell was always in the same community. b) the number of community labels for a given cell as a proportion of the number of days that cell was present in the dataset. This was calculated as the (number of unique community labels / number of days a cell was present). c) stable communities, marked as those which had the same community label for the entire study period.

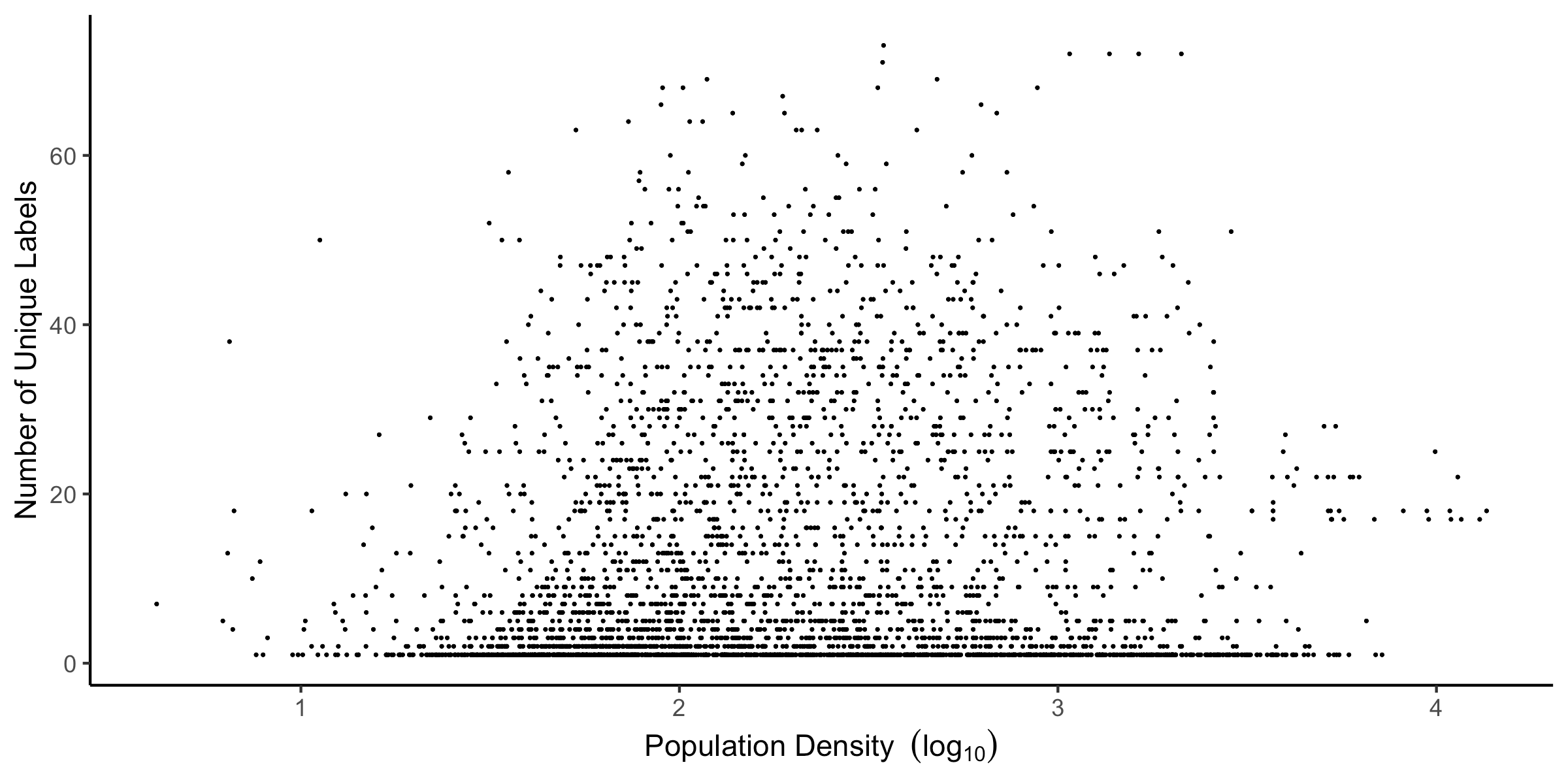

**Supplemental Figure 15. The relationship between community labels and population density.** The relationship the population density of individual cells and the number of community labels assigned during the period. We do not observe a strong association between population density and the number of community labels assigned to individual cells.

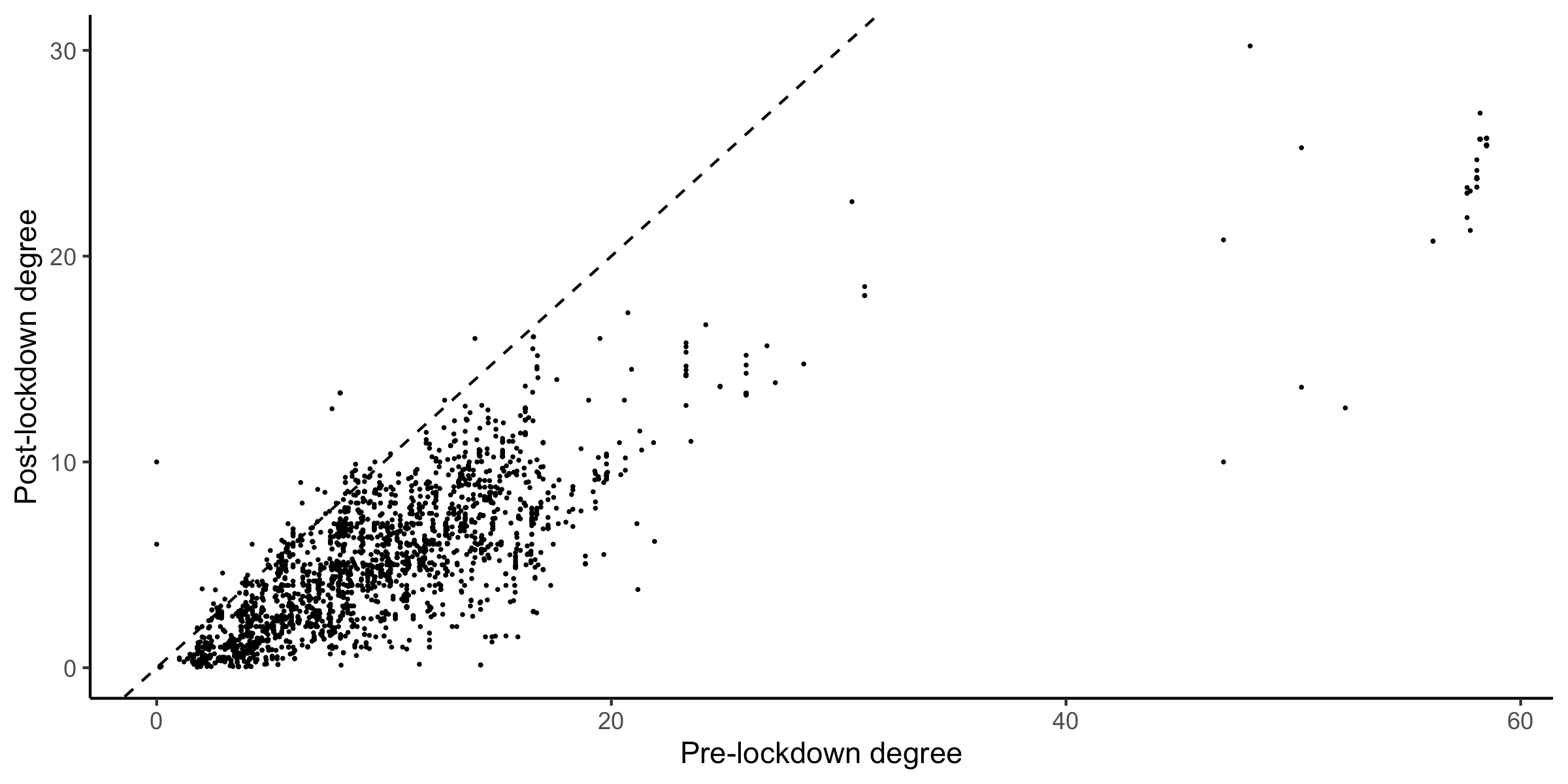

**Supplemental Figure 16. The relationship between community degrees during national interventions.** The total degree of movement communities assigned to individual tiles. We observe a relationship between the degree of communities before and during national interventions, indicating that highly connected communities remained highly connected during the period of national interventions.

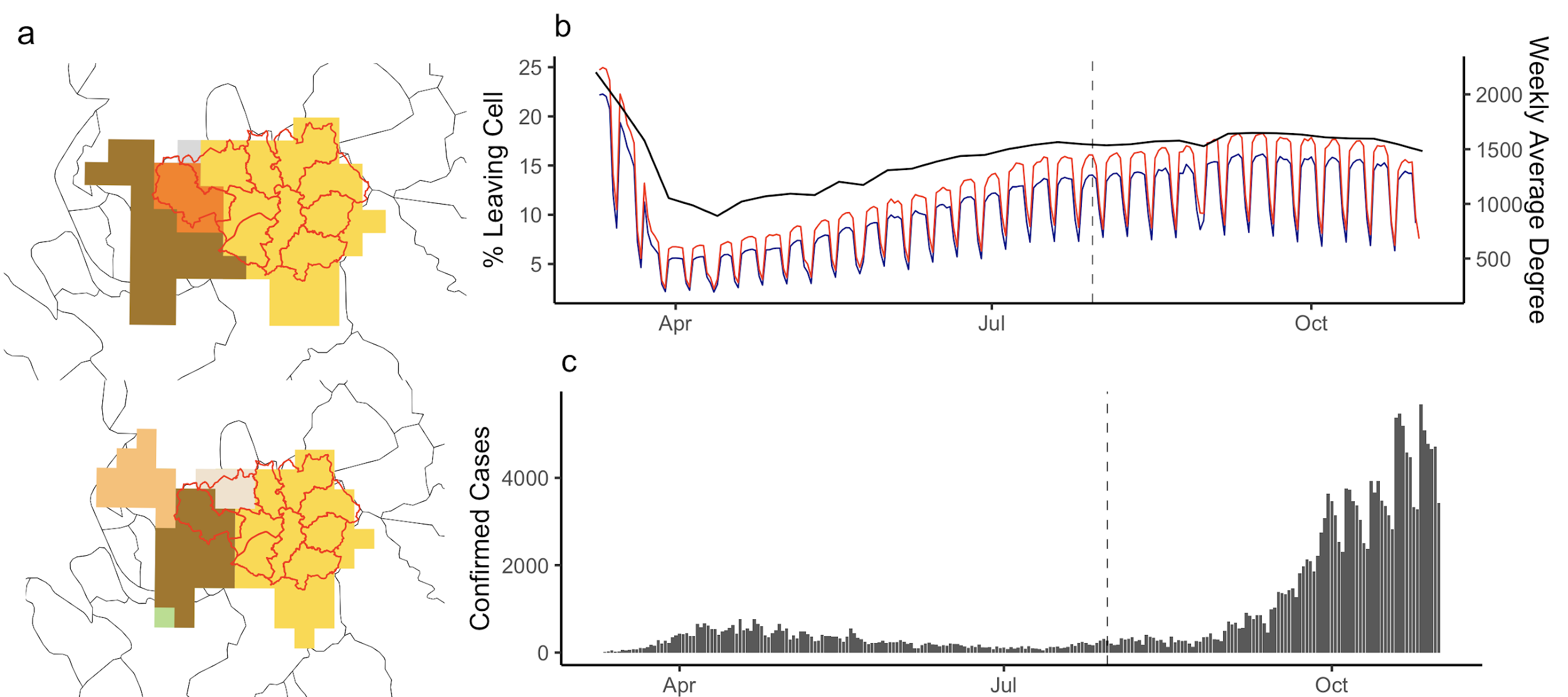

**Supplemental Figure 17. Communities connected to Manchester local area restrictions.** The daily percentage of users travelling between cells in the intervention area and the connected community, and the weekly average degree of intervention cells (a). Confirmed cases in the intervention area (b). Changes in the community structure before and after the introduction of local intervention (c).

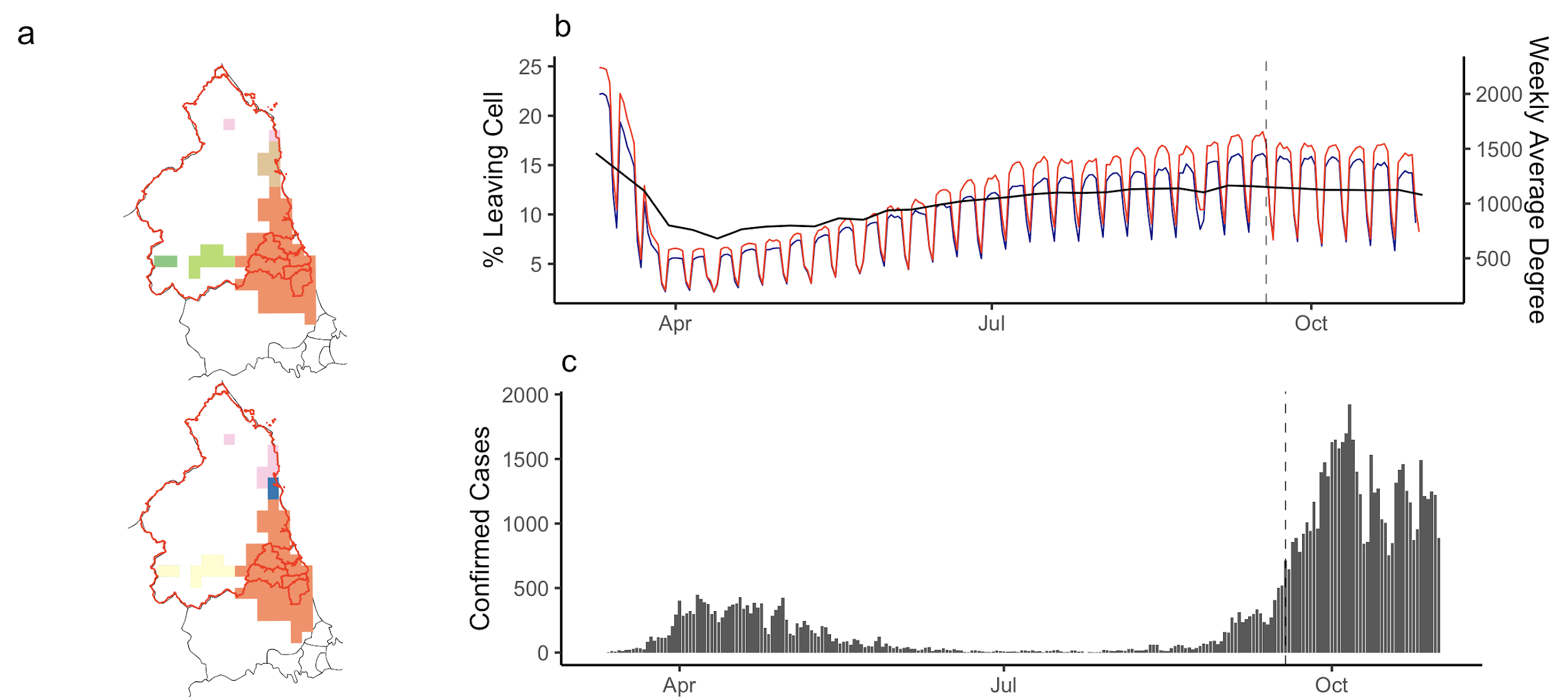

**Supplemental Figure 18. Communities connected to North East England local area restrictions.** The daily percentage of users travelling between cells in the intervention area and the connected community, and the weekly average degree of intervention cells (a). Confirmed cases in the intervention area (b). Changes in the community structure before and after the introduction of local intervention (c).

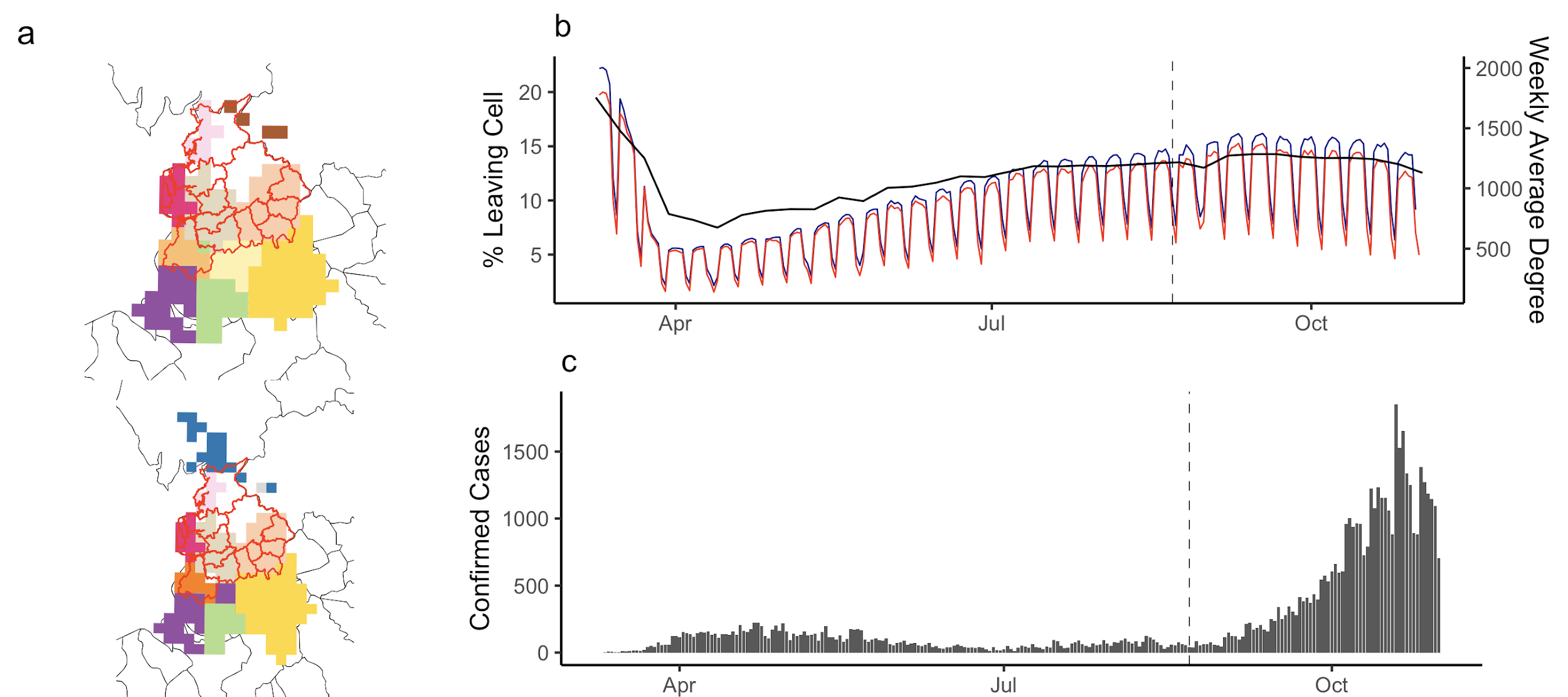

**Supplemental Figure 19. Communities connected to North West England local area restrictions.** The daily percentage of users travelling between cells in the intervention area and the connected community, and the weekly average degree of intervention cells (a). Confirmed cases in the intervention area (b). Changes in the community structure before and after the introduction of local intervention (c).
